# Validation of a remote and fully automated story recall task to assess for early cognitive impairment in older adults: a longitudinal case-control observational study

**DOI:** 10.1101/2021.10.12.21264879

**Authors:** Caroline Skirrow, Marton Meszaros, Udeepa Meepegama, Raphael Lenain, Kathryn V. Papp, Jack Weston, Emil Fristed

## Abstract

**BACKGROUND:** Story recall is a simple and sensitive cognitive test commonly used to measure changes in episodic memory function in early Alzheimer’s disease (AD). Recent advances in digital technology and natural language processing methods make this test a candidate for automated administration and scoring. Convenient and low-burden daily assessments may provide more reliable data than one-off lengthy assessments and be suitable for longer-term disease monitoring.

**OBJECTIVES:** Develop and validate a remote and fully automated story recall task, suitable for longitudinal assessment, in a population of older adults with and without mild cognitive impairment (MCI) or mild AD.

**METHODS:** Participants from AMYPRED-UK (NCT04828122) and AMYPRED-US (NCT04928976) studies were asked to complete optional daily self-administered assessments remotely on their smart devices over 7-8 days. Assessments included immediate and delayed recall of three stories from the Automatic Story Recall Task (ASRT), a test with multiple parallel stimuli (18 short stories, and 18 long stories) balanced for key linguistic and discourse metrics. Verbal responses were recorded and securely transferred from participants’ personal devices, and automatically transcribed and scored using text similarity metrics between the source text and retelling to derive a generalised matching score (G-match). Adherence and task performance differences were examined with logistic mixed models and linear mixed models, respectively. Correlational analysis examined parallel forms reliability of ASRTs, and convergent validity with established cognitive tests (Logical Memory Test, and Preclinical Alzheimer’s Cognitive Composite with semantic processing (PACC5)). Acceptability and usability data were obtained via remotely administered questionnaire.

**RESULTS:** Out of 200 participants recruited into the AMYPRED studies, a total of 151 participants (75.5%, 78 cognitively unimpaired (CU), 73 MCI/mild AD) engaged in optional remote assessments. In these participants, adherence to daily assessment was moderate, did not decline over time, but was higher in cognitively unimpaired participants (66% MCI/mild AD and 78% CU participants completed at least one ASRT story per day). Participants reported favourable task usability: few technical problems, that the application was easy to use, and the tasks were broadly interesting. Task performance improved modestly across the week and was better for immediate recall. G-match scores were lower in participants with MCI/mild AD. Parallel forms reliability of ASRTs were moderate to strong for immediate recall (mean rho=0.73), and delayed recall (mean rho=0.73). ASRTs showed moderate convergent validity with established cognitive tests.

**CONCLUSIONS:** The unsupervised, self-administered ASRT task is sensitive to cognitive impairments in MCI/mild AD. The task shows good usability, high parallel forms reliability and convergent validity with established cognitive tests. Remote, low cost, low burden and automatically scored speech assessments could be used to support diagnostic screening, healthcare and treatment monitoring.

## 1. INTRODUCTION

With the first disease-modifying treatment for Alzheimer’s disease (AD) now available (1), there is an increased need for broader screening and improved monitoring of disease progression and treatment response. Cognitive assessments are currently some of the least invasive, most cost-effective measures available for screening for AD and related impairments. Furthermore, they are supported for use as endpoints of treatment efficacy early in Alzheimer’s disease by key regulatory bodies, including the US Food and Drug Administration (FDA) (2) and the European Medicines Agency (EMA) (3).

However, many cognitive assessments are lengthy, require trained personnel to administer and score and offer few parallel test variants, making them susceptible to practice effects. More importantly, test performance is measurably influenced by a range of state factors, such as sleep (4), exercise (5), mood (6) and stress (7). This variation can give the inaccurate impression of improvement or decline over time (8). Higher frequency sampling can generate more stable and reliable estimates of constructs of interest by controlling for state effects (9) and delineating short-term cognitive fluctuations from longer term changes associated with treatment response and disease progression (8).

Story recall is a cognitive testing paradigm used to assess verbal episodic memory and commonly used to track AD related decline (10–14). Story recall is impaired in Alzheimer’s dementia (15), shows variable differentiation of individuals with mild cognitive impairment (MCI) from those that are cognitively unimpaired (16) and predicts progression from MCI to Alzheimer’s dementia (17).

Most story recall tests are administered in person and scored manually, but research shows that scoring can be fully automated using natural language processing technologies (18). This suggests that story recall tests could be administered in clinic at lower cost and with reduced clinician time burden. Moreover, these tests may be suitable for use in remote assessment, provided that they are properly developed, validated and that test administration can be automated.

Although remote digital assessments are not new, the SARS-CoV-2 global pandemic accelerated the need to adopt remote or hybrid clinical assessment or research methods (19, 20). Alongside advances in technology and connectivity, this has led to a growing appetite for using personal digital devices to collect clinically informative data. Beyond this, digital health technologies can enhance inclusivity, improving access for people who experience mobility problems or those with financial, geographical or time restrictions (21). The continued drive towards remote assessment may be particularly important in older adults who are at substantially increased risk in the pandemic (22). Whilst holding promise for improving convenience and access, there are concerns around whether digital assessment methods are particularly challenging in this population, in particular for those with dementia or milder forms of cognitive impairment (23).

The current study describes the Automatic Story Recall Task (ASRT), a remote, self-administered and automatically scored test developed for repeated cognitive assessment, opening up opportunities for much more nuanced longitudinal data analysis. Test characteristics were examined in participants who are cognitively unimpaired, have mild cognitive impairment (MCI) or have mild AD, assessed repeatedly over one week. The current study examines: (1) acceptability of remote ASRT assessment; (2) adherence to daily ASRT assessments; (3) parallel forms reliability; (4) convergent validity with cognitive and clinical assessments; (5) task performance characteristics; and (6) the effect of daily internal state factors.

## 2. METHODS

### 2.1 Recruitment

Participants were recruited from November 2020-August 2021 in the UK (London/Guildford, Plymouth, and Birmingham), and the USA (Santa Ana, California). Subjects were enrolled if they were cognitively unimpaired (CU) or diagnosed with MCI in the prior 5 years. In the UK study, participants diagnosed with mild AD in the last 5 years were also included. MCI due to AD and mild AD diagnoses were made following National Institute of Aging-Alzheimer’s Association core clinical criteria (24). Subjects were approached if they had undergone a prior amyloid beta PET scan or CSF test (confirmed amyloid beta negative within 30 months or amyloid beta positive within 60 months). Eligibility was established by screening via video call using a secure Zoom link (UK study) or in-clinic assessment (US study), during which the Mini-Mental State Exam (MMSE) (25) was administered. For remote administration no controls for potential environmental prompts to orientation questions (calendars, clocks, watches, etc.) were implemented.

Inclusion criteria comprised age 50-85; MMSE raw score of 23-30 for participants with MCI or Mild AD, 26-30 for CU; cognitively unimpaired or clinical diagnosis of MCI/mild AD made in previous 5 years; English as a first language; availability of a study partner for Clinical Dementia Rating scale (CDR) (26) semi-structured interview; access and ability to use a smartphone running an operation system of Android 7 or above, or iOS 11 or above.

Exclusion criteria: current diagnosis of general anxiety disorder; recent (6-month) history of unstable psychiatric illness; history of stroke within the past 2 years or a documented history of transient ischaemic attack or unexplained loss of consciousness in the last 12 months. Participants treated with medications for symptoms related to AD were required to be stable in these medications for at least 8 weeks before study entry and throughout the study. Participants with a current diagnosis of major depressive disorder (MDD) (UK) or those with current or a 2-year history of MDD (US) were excluded.

### 2.2 Ethics statement

This research was approved by Institutional Review Boards in the relevant research authorities (UK REC reference: 20/WM/0116; US IRB reference: 8460-JGDuffy). Informed consent was taken at the study site (US) or electronically in accordance with HRA guidelines (UK). Studies are registered on clinicaltrials.gov (NCT04828122, NCT04928976).

### 2.3 Procedure

#### 2.3.1 Clinical assessments

Participants completed clinical assessments via a secure Zoom link (UK) or in-clinic (US), completed with a trained psychometrician. Tests reported in the current study are described in detail below.

The Wechsler Logical Memory test (LM) (27) evaluates free recall of a story according to 25 pre-defined information units (IUs: a metric quantifying the amount of information recalled (28)), immediately after presentation, and after a 30-minute delay. Paraphrased answers were accepted and scoring was completed manually according to the instructions and in alignment with the WMS administration and scoring guidelines. Immediate and delayed recall scores were obtained.

Cognitive tests incorporated in the Preclinical Alzheimer’s Cognitive Composite with semantic processing (PACC5) were administered. Tests were manually scored and a mean z-score was calculated as described previously (11). The composite includes summary scores from five measures: (1) the MMSE (25), a global cognitive screening test; (2) the Logical Memory Delayed Recall (27), a delayed story recall test; (3) Digit-Symbol Coding (29), a symbol substitution test; (4) the sum of free and total recall from the Free and Cued Selective Reminding Test (30), a multimodal associative memory test; and (5) Category Fluency (animals, vegetables, fruits), a semantic memory test.

The CDR (26), a semi-structured interview assessing severity of cognitive symptoms of dementia, was completed with the participant and their study partner and scored based on the CDR Sum of Boxes (CDR-SB) scales. The examiner was not blinded to other assessments administered. In the US study, where participants had completed subtests of the PACC5 or CDR assessments within one month prior to the study visit, tests were not re-administered but the recent historical test results were used.

Participants completed the Automatic Story Recall Task (ASRT), a task constructed to elicit naturalistic speech within a closed domain. Pre-recorded ASRTs are presented at a steady reading rate (approximately 140 words per minute) by a British male speaker. Parallel stimuli include 18 short stories (mean of 30 IUs and 119 words per story) and 18 long stories (mean of 60 IUs and 224 words per story). Task characteristics are presented in the multimedia appendix table S1, showing that stories incorporate a range of themes and are balanced for key linguistic and discourse metrics. During clinical assessments, three long ASRT stories were administered consecutively. After each story was presented, participants were asked to immediately retell the story in as much detail as they could remember. Recall of the same stories, in the same order, was tested again after a delay.

During clinical assessments, participants were supported with installing the Novoic mobile application (‘the app’) on their own smartphone device and were shown how to use it. Participants were reimbursed for their participation at the end of the study visit and prior to remote assessments (£65 for UK participants $75 for US participants).

#### 2.3.2 Remote assessments

Participants were encouraged to complete optional unsupervised self-assessments (under 30 mins in length) on the app daily for up to eight days following the study visit. Remote assessments included the Automatic Story Recall Tasks (ASRTs), administered daily, in threes (triplets) and at the beginning of each remote assessment session. The three ASRTs administered on the first day of remote assessment were identical and repeated stories from those administered in clinical supervised assessment on the prior day. The remainder of ASRT stories, presented from day 2 of remote assessment onwards, were novel and administered only once.

After each story was presented, participants were asked to immediately retell the story in as much detail as they could remember. Recall of the same stories, in the same order, was tested again after a delay. The schedule included delayed recall after completion of all immediate recalls, or after completion of brief distractor tasks (category or verbal fluency), with test method varying by day (the multimedia appendix table S2). Task responses were recorded on participants’ personal smart devices and automatically uploaded to a secure server.

Due to participant feedback regarding high burden, the assessment schedule was changed partway through the study. The new schedule favoured shorter stories and reduced the number of additional assessments following ASRTs (not reported here). Simultaneously, the number of days of remote assessment was increased from seven to eight days to spread out assessments. Details are provided in the multimedia appendix table S2.

Daily state effects were assessed at the end of each remote assessment via a four-item self-report questionnaire asking how participants were feeling that day (current mood, sleep, mind-wandering and effort). App and task usability was assessed via a self-report questionnaire on day 2 (initial assessment schedule) or day 5 (revised assessment schedule). Participants reported technical difficulties experienced during assessments, whether technical difficulties prevented them from completing the assessments, how easy and how interesting it was to complete the assessments. Questionnaires are shown in the multimedia appendix tables S3 and S4.

### 2.4 Statistical analysis

Stories were transcribed using an out-of-the-box automatic speech recognition (ASR) system, followed by automated textual analysis completed using a generalized matching score (referred to here for brevity as “G-match”), computed in Python as the weighted sum of the cosine similarity between the embeddings of original ASRT text and the transcribed retellings. G-match provides an index of the proportional recall for each story, with potential scores ranging from 0 to 1 (hypothetically perfect performance). Mean G-match per triplet was also computed.

Further analysis was completed in R v.4.0. Data were assessed for normality, followed by parametric and non-parametric analyses, as appropriate. Adherence was defined as engaging with at least one ASRT story per day. Adherence patterns were examined with logistic regression models, predicting adherence at immediate and delayed recall, in relation to participant group, demographics, assessment day and schedule. A large proportion of participants only completed seven days of remote assessments (table 1), and longitudinal analysis of adherence was therefore limited to assessments on days 1-7. Participants were included as random effects. Demographics (sex, age, years in education), assessment days (1−7), research schedule (schedule 1 or schedule 2) and participant group (CU or MCI/mild AD) were included as fixed factors.

**Table 1:** Participant demographics: Demographic characteristics of cognitively unimpaired and MCI/mild AD participants. MCI: mild cognitive impairment; AD: Alzheimer’s dementia; CU: cognitively unimpaired; N, number; SD, standard deviation; MMSE: Mini-Mental State Exam; UK: United Kingdom; US: United States of America.

|  | Group |  | Statistic and *P*-value |
| --- | --- | --- | --- |
|  | Cognitively unimpaired | MCI/mild AD |  |
| Total sample size | 78 | 73 | - |
| Number female/ male | 47/31 | 41/32 | $\chi^2=0.26$, $p=0.61$ |
| Number of participants from UK/US studies | 66/12 | 51/22 | $\chi^2=4.70$, $P<.03$ |
| Schedule 1 versus schedule 2 | 40/38 | 22/51 | $\chi^2=6.97$, $P=.008$ |
| Amyloid beta negative/positive | 38/40 | 41/32 | $\chi^2=0.84, P=.36$ |
| Mean years in education (SD) | 15.24 (3.37) | 15.06 (2.80) | $r=-0.05, P=.57$ |
| Mean age (SD) | 70.37 (4.35) | 69.58 (7.30) | $r=-0.008, P=.91$ |
| Mean MMSE (SD) | 28.92 (1.15) | 27.00 (2.07) | $r=0.50, P<.001$ |

Parallel forms reliability of ASRTs was examined with pairwise correlational analysis. Only ASRTs administered across both schedules were analysed, maintaining comparable sample sizes across comparisons and allowing for testing within MCI/mild AD and CU subgroups. Convergent validity of these same ASRT stories was examined in relation to the LM, PACC5 and CDR-SB. Analyses were repeated with the mean G-match score per triplet. To improve consistency and comparability of reporting, Spearman’s rank correlation coefficients are reported.

Task performance differences between groups, task administration variations and change over time were modeled using longitudinal linear mixed effects models. Data analysed was restricted to days 2-7 where all assessments were novel and administered to all participants. Analysis included G-match as the response variable, and fixed effects of participant group, remote assessment days (2-7), order (1st, 2nd or 3rd ASRT presented), long or short stories and immediate or delayed recall. Demographics (age, sex, education) were included as additional fixed effects. A random effect of participant with random slope and intercept was specified. Cohen’s d scores for multilevel model objects were calculated using the lme.dscore command in the package EMAtools.

Analyses were repeated with the mean G-match per triplet, with equivalent random and fixed effects specifications, excepting story order, which was not included. Covariation of mean ASRT task performance across triplets with self-reported daily state was then examined, by additionally incorporating fixed effects of self-reported mood, sleep, effort and mind-wandering, into the above model. Assumptions of all regression models were investigated by examining the distribution and patterns of residuals versus fitted values.

## 3. RESULTS

### 3.1 Participants

Two hundred participants, 67 from the US study and 133 from the UK study, were recruited and completed the clinical assessment protocol. One hundred and fifty-one participants (75.5%) completed at least one remote ASRT. Older participants (r=-0.15, *P*=.03), those with lower MMSE scores (r=-0.26, *P*< .001) and those with MCI/mild AD (33/106 MCI (31%) and 16/94 CU (17%); χ^2^=5.36 (DF=1), *P*=.02) were less likely to complete any remote assessments. There were no differences in sex ratio (χ^2^=0.41 (DF=1), *P*=.52), or years in education (r=-0.01 *P*=.87) between participants who contributed at least one remote assessment and those who did not.

Demographic information for participants providing remote data are presented in table 1. In this sample, MCI and CU groups did not differ with respect to age, years in education, sex or amyloid status. The US study included proportionally more participants with MCI. The MCI/mild AD group included a minority of participants with a diagnosis of mild AD (10/73, 13.7%).

### 3.2 Usability

Usability questionnaires were completed by 63% (96/151, n=52 CU, n=44 MCI/mild AD) of participants who completed remote assessments (figure 1). Those completing usability questionnaires (N=96/151) did not differ with respect to education level (r=-0.02, *P*=.78), age (r=-0.12, *P*=.14), or MMSE scores (r=-0.08, *P*=.32) compared to those who engaged in remote assessments but did not complete usability questionnaires (N=55/151). There was also no difference in the male/female ratio (χ^2^=0.11 (DF=1), *P*=.75) or the ratio of CU to MCI/mild AD participants (χ^2^=0.67 (DF=1), *P*=.41) who did and did not complete usability questionnaires.

**Figure 1:**
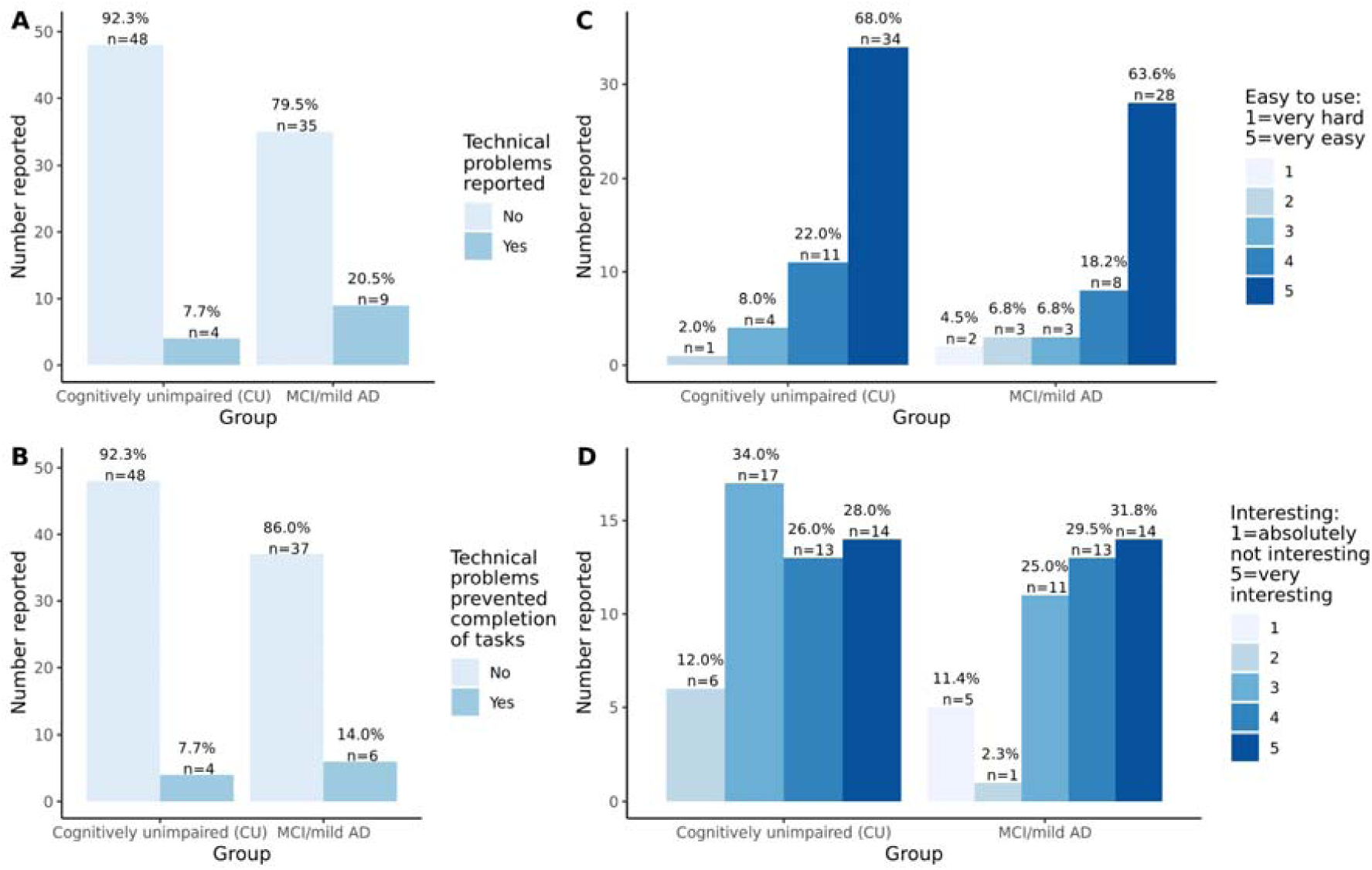
responses to usability questionnaire: A) technical problems reported; B) rate at which technical problems prevented completion of tasks; C) ease of use of application; D) interest in tasks completed.

Participants reported few technical difficulties and most reported that technical difficulties had not prevented them from completing the assessments, with no group differences (χ^2^=3.32 (DF=1), *P*=.07 and χ^2^=0.98 (DF=1), *P*=.32, respectively). Participants overwhelmingly responded that the app was easy to use, and that the task was reasonably interesting, with no group differences (r=-0.08, *P*=.47 and r=-0.04, *P*=.70, respectively).

### 3.3 Adherence

Participants with MCI/mild AD completed fewer remote assessments than CU participants (adherence for immediate recall: 65% versus 78%; delayed recall: 62% versus 77%; figure 2). Group differences were confirmed by mixed logistic regression analyses (immediate recall, estimate=-0.97, *P*=.01; delayed recall estimate =-0.84, *P*=.02). Adherence did not change over assessment days (immediate recall estimate=-0.04, *P*=.34; delayed recall estimate=-0.07, *P*=.11), but lower adherence to delayed recall was seen for the revised test schedule (estimate=-0.86, *P*=.03). Adherence was not associated with sex and education (all *P*>0.2), but younger participants completed more immediate recall assessments (estimates: immediate=-0.07, *P*=.02; delayed=-0.06, *P*=.06).

**Figure 2:**
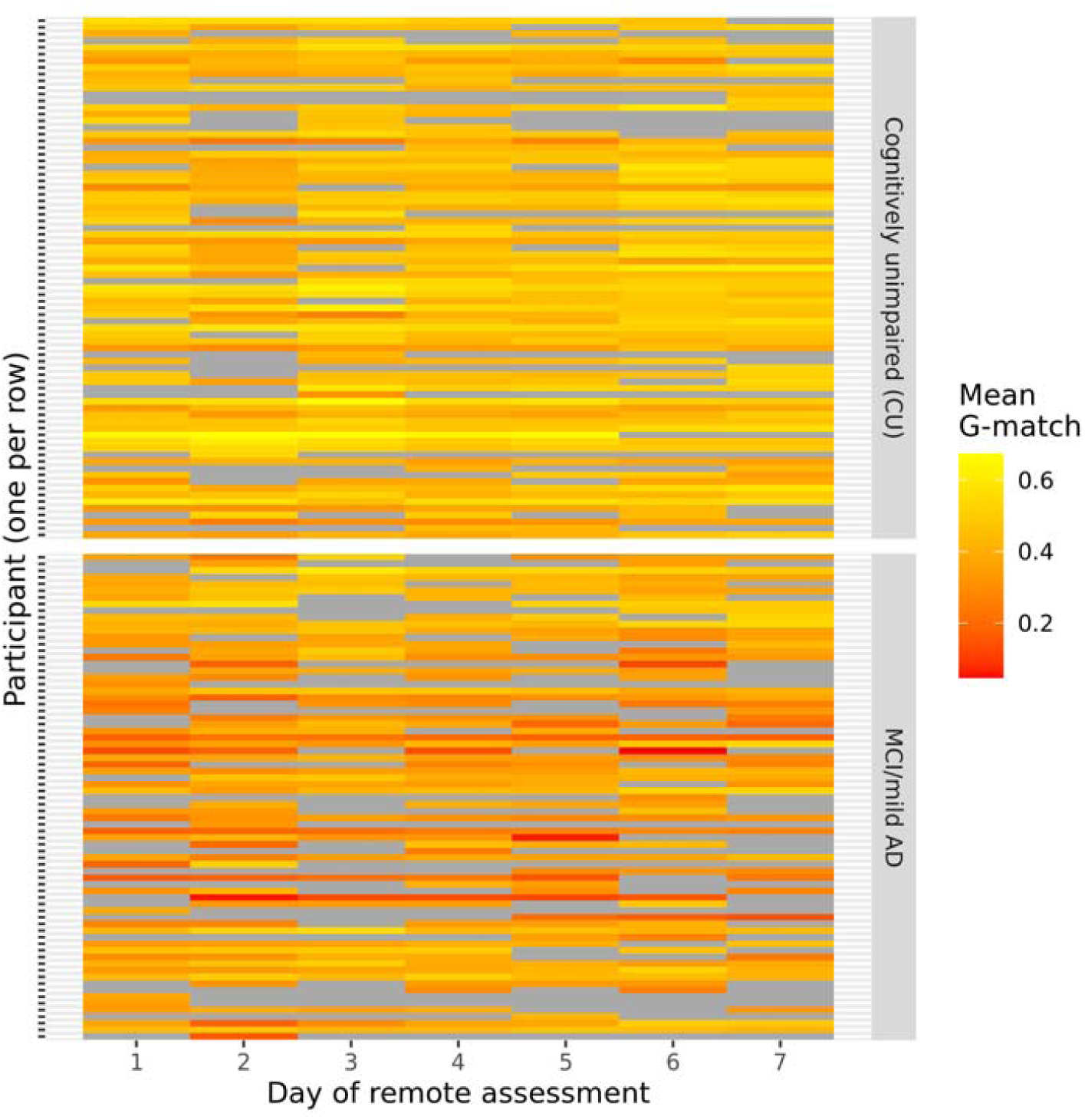
Adherence and task performance heatmap for G-match in immediate recall trials. G-match is an automated measure of recall performance (see methods). Results plotted across individual days of remote assessment for n=151 participants who completed at least one assessment. Each participant is represented by a row, missing data are shown in grey, and mean G-match across ASRT triplets is shown in colour (red=low recall; yellow=high recall).

Figure 2 shows a heatmap of adherence patterns and task performance. In this figure each participant is represented by a row and task response and performance over the days of assessment is shown in coloured blocks along the x-axis. Task performance is shown in colour with red-to-yellow grading representing low-to-high G-match scores. Missing data is shown in grey. The figure reflects the results reported above, with higher adherence in the cognitively unimpaired group, and no clear decline in adherence over the assessment period.

### 3.4 Task characteristics

G-match for ASRTs and triplets showed good psychometric properties. Data generated showed no ceiling or floor effects (fig 3A; multimedia appendix Figure SF1-4). Task performance characteristics are provided in the multimedia appendix tables S5-S7.

**Figure 3:**
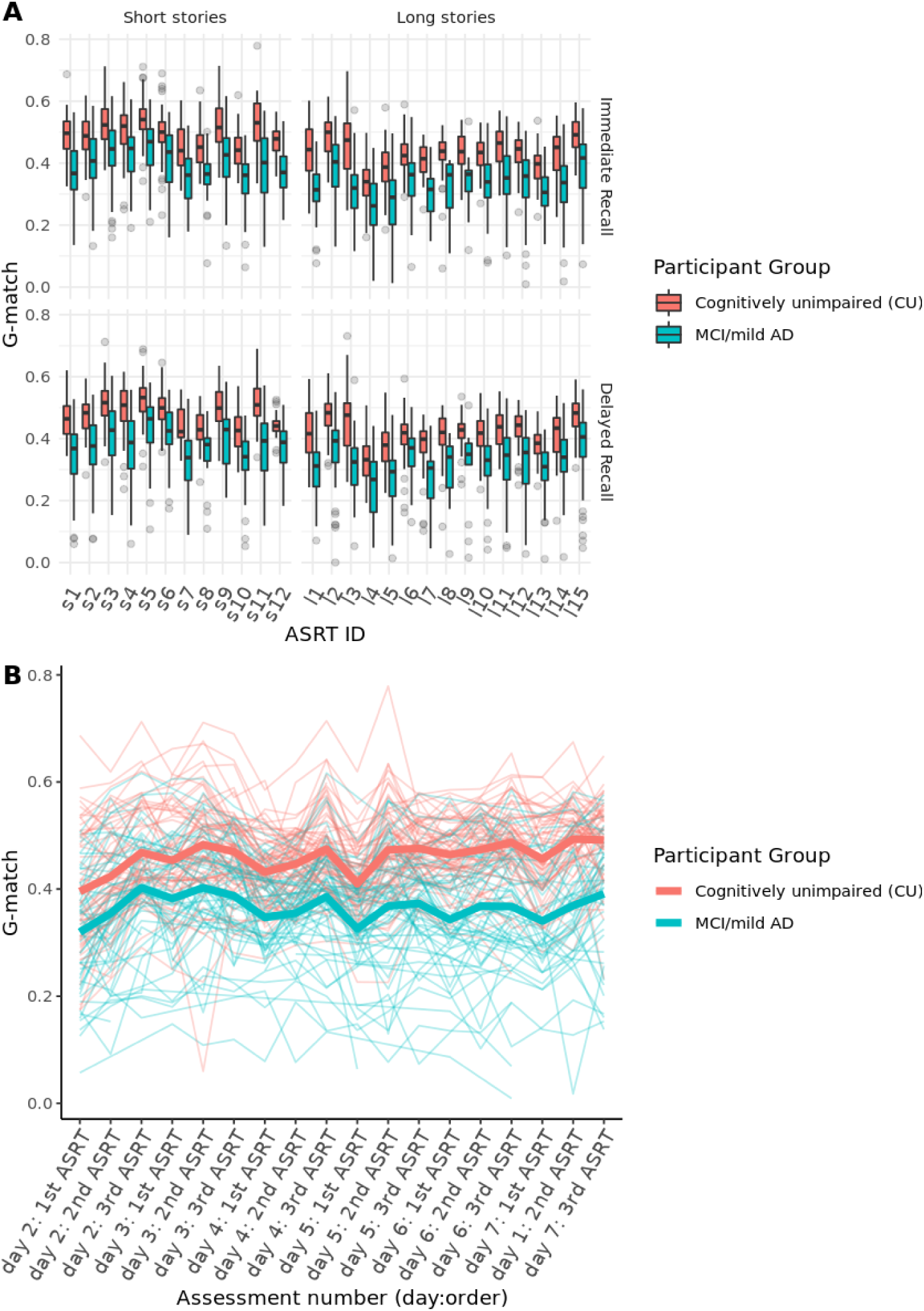
G-match over repeated assessments: A) boxplots of G-match for individual ASRT stories split by short and long story horizontally and by immediate and delayed recall vertically; B) Average G-match (immediate recall) over individual assessment days (2-7, immediate recall), and testing order. Group means for immediate recall in thick lines and individual variability across remote assessment days and testing order with individual trajectories in the paler, thinner lines, showing variability within individuals, across testing days and order of administration. G-match (generalised match) is an automated measure of recall performance (score range 0 (lowest) - 1 (highest)).

### 3.5 Parallel form reliability

Parallel forms reliability for individual ASRT stories at immediate recall are presented in figure 4. Equivalent figures for delayed recall, and separated by clinical group, are presented in the multimedia appendix Figures SF 5-9. Correlation matrices for triplets broken down by immediate and delayed, and clinical groups, are shown in the multimedia appendix Figures SF 10-12.

**Figure 4:**
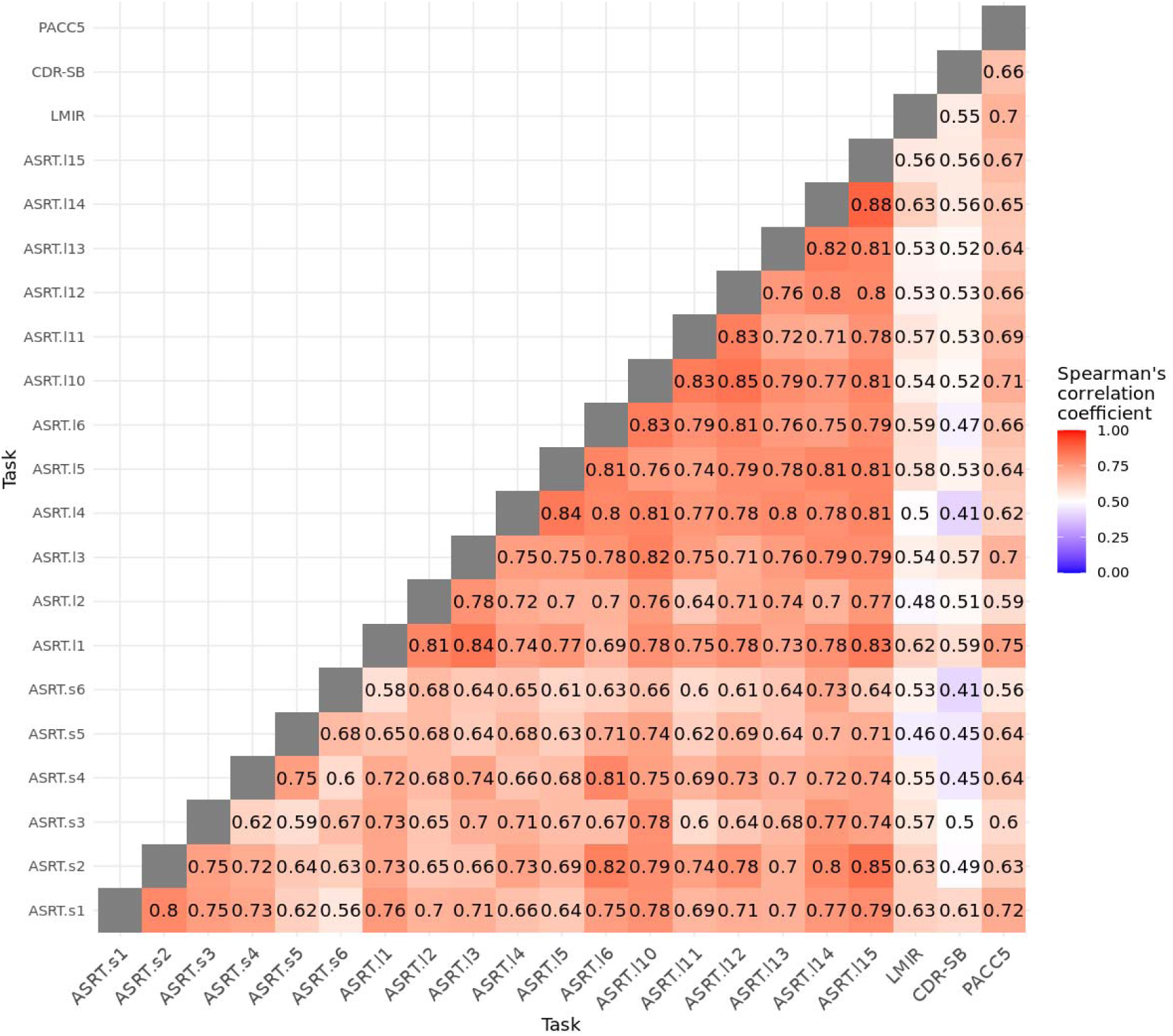
Matrix of pairwise correlation coefficients of test-retest reliability for G-match of individual ASRT stories (immediate recall), and convergent validity with other cognitive and clinical assessments. ASRT stories are denoted with s (for short) and l (for long) followed by the story number (see multimedia appendix table S6). Convergent validity is examined through correlations between ASRT stories with other assessments completed (LMIR, CDR-SB, PACC5). To maintain consistent reporting, the sign of the correlation for the CDR-SB (in which higher scores reflect greater impairment) is reversed. Pairwise correlation coefficients for ASRTs reflect parallel task performance metrics for between n=75-116 participants, and correlation of ASRTs with other cognitive tests for n=92-115, depending on adherence patterns. Abbreviations: ASRT: Automatic Story Recall Task, LMIR: Wechsler Logical Memory Test - Immediate Recall, CDR-SB: Clinical Dementia Rating - Sum of Boxes, PACC5: Preclinical Alzheimer’s Cognitive Composite with semantic processing., G-match: generalised match, an automated measure of recall performance.

Correlation coefficients in the full sample were moderate to strong for immediate recall (rho range=0.56-0.88, mean=0.73), and remained so after restricting analyses to MCI/mild AD (rho range=0.31-0.87, mean=0.65) and CU (rho range 0.39-0.85, mean=0.65). Similarly, correlations between parallel ASRT stories were moderate-high for delayed recall (full sample: rho range=0.54-0.86, mean=0.73), and remained so when restricting analyses to MCI/mild AD (rho range=0.37-0.88, mean=0.65) and CU participants (rho range=0.32-0.83, mean=0.64).

Test-retest reliability was higher when examined for mean scores obtained across triplets (immediate; rho range=0.77-0.88, mean=0.83; delayed: rho range=0.76-0.89, mean=0.85), remaining consistently high in MCI (immediate; rho range=0.57-0.88, mean=0.73; delayed: rho range=0.60-0.89, mean=0.75) and CU subgroups (immediate; rho range=0.67-0.83, mean=0.76; delayed: rho range=0.68-0.85, mean=0.77).

### 3.6 Convergent validity

ASRT task performance correlated moderately with other cognitive and clinical measures (LM, CDR-SB and PACC5) in the full sample across both immediate and delayed recalls (figure 4). Mean correlation coefficients between immediate recall ASRTs with LM-Immediate recall (LMIR), PACC5 and CDR-SB, were rho=0.56, 0.65, and 0.51, respectively. Mean correlation coefficients between ASRTs with LM-delayed recall (LMDR), PACC5 and CDR-SB were rho=0.54, 0.66, and 0.50, respectively. Analysis results and figures for delayed recall, and results separated by participant group are provided in the multimedia appendix Figures SF 5-9). Correlation coefficients remained in the moderate range after restricting analyses to participants with MCI/mild AD but were typically lower in CU participants. Correlations between ASRT triplets and other cognitive tests are provided in the multimedia appendix figures SF10-12.

### 3.7 Task performance

Longitudinal mixed models are presented in table 2, with similar results for individual ASRTs and triplets. Task performance improved across the week, with a modest linear daily improvement estimate in G-match by assessment day. There was an effect of group with lower scores in the MCI/mild AD group for both individual stories and triplets, with an equivalent effect size of Cohen’s d=1.54. G-match was higher for immediate recall, shorter stories, and higher for the latter ASRTs administered within each triplet. Demographics were not associated with task performance. Longitudinal data are displayed visually in figure 3B, showing within and between-subject variability.

**Table 2:**
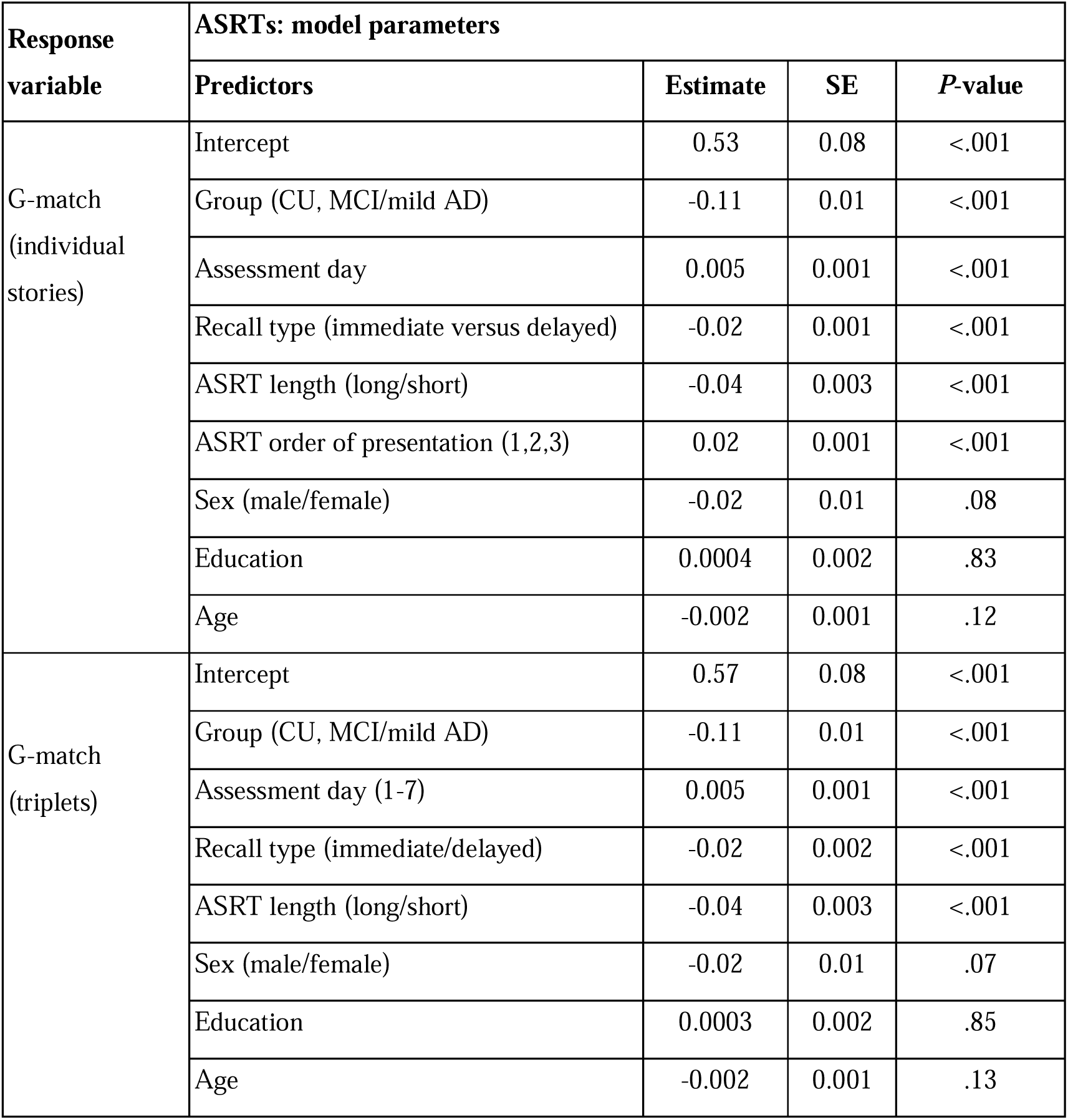
Effects of task characteristics, participant group, and demographics on task performance metrics as estimated by longitudinal mixed models: MCI: mild cognitive impairment; AD: Alzheimer’s dementia; CU cognitively unimpaired; G-match: Generalized match

| **Response variable** | **ASRTs: model parameters** |  |  |  |
| --- | --- | --- | --- | --- |
|  | **Predictors** | **Estimate** | **SE** | **P-value** |
| G-match (individual stories) | Intercept | 0.53 | 0.08 | <.001 |
|  | Group (CU, MCI/mild AD) | -0.11 | 0.01 | <.001 |
|  | Assessment day | 0.005 | 0.001 | <.001 |
|  | Recall type (immediate versus delayed) | -0.02 | 0.001 | <.001 |
|  | ASRT length (long/short) | -0.04 | 0.003 | <.001 |
|  | ASRT order of presentation (1,2,3) | 0.02 | 0.001 | <.001 |
|  | Sex (male/female) | -0.02 | 0.01 | .08 |
|  | Education | 0.0004 | 0.002 | .83 |
|  | Age | -0.002 | 0.001 | .12 |
| G-match (triplets) | Intercept | 0.57 | 0.08 | <.001 |
|  | Group (CU, MCI/mild AD) | -0.11 | 0.01 | <.001 |
|  | Assessment day (1-7) | 0.005 | 0.001 | <.001 |
|  | Recall type (immediate/delayed) | -0.02 | 0.002 | <.001 |
|  | ASRT length (long/short) | -0.04 | 0.003 | <.001 |
|  | Sex (male/female) | -0.02 | 0.01 | .07 |
|  | Education | 0.0003 | 0.002 | .85 |
|  | Age | -0.002 | 0.001 | .13 |

After incorporating self-report assessments into the mixed model predicting G-match for triplets, models revealed a significant effect of mood (estimate=0.007 (SE=0.002), *P*<.001) and mind-wandering (estimate=-0.007 (SE=0.002), *P*<.001), with better daily mood and lower mind-wandering associated with better daily task performance.

## 4. DISCUSSION

The current study indicates that daily unsupervised and self-administered speech-based testing is acceptable and feasible in older participants with and without cognitive impairment. Participants engaged in daily optional assessments with moderate levels of adherence, and no observable reduction in adherence levels over the week-long period of assessment. Participants experienced few technical problems and reported that the tests were easy to use and reasonably interesting.

Results indicate that remote automatic test administration and auto-scoring of story recall can provide sensitive cognitive measurement in at-risk populations. ASRT generalised match (G-match), an automatically scored measure of proportional recall, showed consistent differences in task performance between cognitively healthy participants and those with MCI/mild AD. Separation in task performance between diagnostic groups was consistent across the assessment period, and across individual ASRT stories (figure 3), showing a strong effect size for differentiating cognitively unimpaired participants from those with MCI/mild AD (Cohen’s d=1.54), whilst controlling for age, education and sex. The equivalent area under the receiver operating characteristic curve (AUC) is 0.86, based on previously published equivalence tables (31).

The ASRTs discrimination between clinical groups reported here outperforms those previously reported for differentiating cognitively unimpaired individuals from those with MCI using traditional cognitive tests administered in person and in clinic, such as the MMSE (Cohen’s d=0.69) and the 6-Item cognitive impairment test (d=0.65), and Addenbrooke’s Cognitive Examination-Revised (d=0.73), albeit with similar results reported previously for the Montreal Cognitive Assessment battery (d=1.45) (32). Similarly, evidence suggests that the task performs better for differentiating cognitively unimpaired individuals from those with MCI on traditional story recall tests such as the Logical memory-II test, with reported AUCs of 0.61-0.72 (16). The test also performs well in comparison to the Cogstate brief battery, when remotely administered and unsupervised, where effect sizes for differences between MCI and cognitively unimpaired groups in subtests range from Cohen’s d=0.22-0.62 (33).

Although the mixed clinical group examined in the current analyses initially limits direct comparison with these previously published metrics, the mild AD group was only a small proportion of (10/73, 13.7%) of those contributing to the MCI/mild AD group. After excluding participants with mild AD from linear mixed model analysis, this yields an effect size of Cohen’s d=1.45 (equivalent AUC=0.85 (31)) for the difference between cognitively unimpaired and MCI participants.

ASRT stimuli are carefully designed and balanced for key linguistic and discourse metrics, including number of words, number of sentences, number of dependent clauses, mean sentence length, and ratio of dependent clauses to t-units. This balancing of the stimuli is also reflected in good parallel forms reliability between ASRT parallel stimuli, which is consistently high across immediate and delayed recall, and remains so within clinical subgroups (MCI/mild AD or cognitively unimpaired).

Repeated exposure to the test stimuli may lead to unwanted practice effects that reduce the validity of the test as a measure of new learning, with research also showing differential practice effects across clinical diagnostic groups for tests such as list learning tests and the Logical memory test (34). Practice effects may be particularly important to consider where the same story recall stimuli are used repeatedly in longitudinal research or clinical monitoring, or for diagnostic thresholding as cut-offs for research studies or clinical trials (16,35). With certain exceptions such as the Craft stories (36), available story recall tests typically have a limited range of parallel forms.

Whilst alternate test variants can help to reduce practice effects, they do not completely correct for retesting, which can be modified by repeated exposure to the task and greater familiarity with the test structure or method (37). In the current study, despite novel stimuli being presented during each assessment, test scores improved modestly during the week, indicating that increased familiarity with the app, testing procedure and/or test structure resulted in a subtle improvement over time. Task performance improvements over the weeklong assessment period were modest (with an estimated daily change in G-match of only 0.36% of the initial intercept estimate value).

ASRT tests correlated moderately with a well-established test of verbal episodic memory, cognitive composites and clinician reported outcomes, indicating acceptable convergent validity, and with results comparable to, or better than, other studies of computerised or unsupervised remote assessments (38–40). Correlations with the Logical Memory test and clinician reported outcomes were in the moderate range, with lower correlation coefficients linked to test invariance due to ceiling or floor-level performance on these assessments in cognitively unimpaired individuals.

Task performance also varied with aspects of study design, with stories administered later in triplets delivering more comprehensive recall. These effects appear to lead to greater variation between individual ASRT stories but are averaged out when G-match is examined across story triplets.

Analysis of story triplets shows higher parallel forms reliability between ASRTs administered and analysed in threes, albeit with broadly unchanged measurable differences in group performance. Task performance as measured with G-match was typically higher for shorter stories, indicating that responses more comprehensively covered the story source text where participants were asked to recall less material.

The current study also shows within-subject variation in task performance, in part reflecting measured effects of state factors on cognitive performance, in particular daily mood and effort. Variation from within-subject differences can make it challenging to differentiate clinical change from measurement error (9), and higher-frequency assessments carried out longitudinally can help to generate more reliable estimates of cognitive function and change. Repeated measurements allow these state effects to be concurrently measured and included or controlled for in longitudinal analyses.

### 4.1 Limitations

Whilst our study shows that a high proportion of participants engaged in optional remote assessments, older and more cognitively impaired participants were less likely to contribute to this part of the study. In response to participants’ and study centres’ feedback of high participant burden of the initial test schedule, the testing schedule was altered in the middle of the study to reduce burden, thereby limiting the data available for certain ASRT test variants. Adherence to remote assessment also varied by clinical group, with a greater number of assessments completed by cognitively unimpaired and younger participants. Assessment under supervision, either in clinic or during a telemedicine visit, allowing for provision of additional support where required, could be better suited to more impaired subjects.

The design of the study makes it difficult to differentiate between the effects of individual stories themselves (i.e. which ASRT story was used) and effects of study design, such as test order or day of assessment. Future studies may benefit from adopting a randomised design, with ASRTs randomly selected and allocated to different testing instances, to derive test performance metrics independent of these additional confounders. For longitudinal studies, either short or long stories should be adopted to improve consistency of test scores over time and help to better characterise cognitive change.

Participants included in the current study constitute a select sample. The sample was selected to exclude concurrent neurological and mental health conditions. They were recruited from prior clinical trials completed in the US and the UK and reflect a group of individuals who are actively engaged in clinical research. The participants are lacking in racial diversity (with the majority of the sample identifying as white, and with only 2.6% (N=4) of Asian, Black, African or African American background) Replication is now needed in more clinically and demographically heterogeneous samples.

### 4.2 Overview and future directions

The recent FDA approval for the first disease modifying treatment for people at risk of developing AD highlights the importance of adequate screening and early detection, as well as the importance of monitoring treatment response. Briefer, convenient and lower-burden daily assessments may provide more reliable data to evaluate disease progression or treatment response than one-off lengthy assessments (9). Brief digital assessments, completed at home, and repeatable over time could improve access to AD screening as compared to current clinical standards, which typically required clinical visits and extensive neuropsychological assessment.

The current study shows that brief, remotely administered and automatically scored ASRTs are sensitive to early cognitive impairments commonly identified through more extensive clinical assessment. The tests show good properties for repeated administration, and convergent validity with established tests of episodic memory, cognitive composites and clinician reported outcomes (CDR-SB). The test shows good acceptability and usability in older adults with and without cognitive impairment. Furthermore, due to the automatic administration and scoring of ASRTs, this test presents a minimal administrative burden, requiring no trained personnel or specialist equipment.

Speech is instrumental to daily functioning, and a natural response modality for participants to use in response to current smart devices, such as smartphones. Speech responses are also a common component of cognitive tests, however data generated in these tests, including those reported in this study, often relate to simple pass/fail characteristics of response accuracy. New metrics using audio- and text-based AI models to target other changes measurable in speech data (acoustic (41,42), semantic (43–46), linguistic (42)) in early-stage Alzheimer’s disease could further leverage the information content of ASRTs, developing a new class of powerful, fully automated speech biomarkers.

## Supporting information

Multimedia

## Data Availability

Speech data is identifying and cannot be shared, but all quantitative data produced in the present study are available upon reasonable request to the authors.

## Acknowledgments

We are extremely grateful to our participants who took part in the study and their families/carers who supported their participation. We also thank the study sites and their scientific and research team for recruitment, study coordination, conducting interviews and data collection efforts.

## Author contributions

The study protocol was designed by EF, MM, and JW. The research study and data collection were coordinated by MM. Analyses were completed by CS, UM, and EF. The first draft of the manuscript was completed by CS and EF. All authors contributed to the revision of the manuscript.

## Abbreviations

AD: Alzheimer’s disease
AI: Artificial intelligence
ASRT: Automatic Story Recall Test
AUC: Area under the curve
CDR-SB: Clinical Dementia Rating - Sum of Boxes
CU: Cognitively unimpaired
G-match: Generalised match
LM: Logical memory test
LMIR: Logical memory immediate recall
LMDR: Logical memory delayed recall
MCI: Mild cognitive impairment
MMSE: Mini-Mental State Exam
N: Number
PACC5: Preclinical Alzheimer’s Cognitive Composite with semantic processing
ROC: Receiver Operating Characteristic
SD: Standard deviation
UK: United Kingdom
USA: United States of America

