## Supplementary material for "Validation of a remote and fully automated story recall task to assess for early cognitive impairment in older adults: a longitudinal case-control observational study": Multimedia

**Multimedia appendix**

### Supplementary Table S1: ASRT task characteristics

| **ASRT ID** | **Length** | **Story name** | **Number of** | | | | **Mean sentence length** | **Ratio of dependent clauses to T-units** |
| --- | --- | --- | --- | --- | --- | --- | --- | --- |
|  |  |  | dependent clauses | T units | words | sentences |  |  |
| l1 | Long | No ordinary thief | 8 | 32 | 201 | 14 | 14.36 | 0.25 |
| l2 | Long | The music lover | 8 | 36 | 210 | 14 | 15.00 | 0.22 |
| l3 | Long | The walled city | 8 | 32 | 201 | 14 | 14.36 | 0.25 |
| l4 | Long | The art competition | 7 | 33 | 223 | 15 | 14.87 | 0.21 |
| l5 | Long | The Big Beetle | 7 | 34 | 220 | 14 | 15.71 | 0.21 |
| l6 | Long | A soldier returns | 10 | 41 | 223 | 15 | 14.87 | 0.24 |
| l7 | Long | The voice of God | 7 | 39 | 224 | 16 | 14.00 | 0.18 |
| l8 | Long | The silent nun | 11 | 41 | 225 | 16 | 14.06 | 0.27 |
| l9 | Long | The king and the doctor | 9 | 31 | 220 | 16 | 13.75 | 0.29 |
| l10 | Long | Lost on an Island | 11 | 34 | 222 | 14 | 15.86 | 0.32 |
| l11 | Long | Treasure of the Pyramids | 10 | 35 | 217 | 14 | 15.50 | 0.29 |
| l12 | Long | The dry swimmer | 10 | 41 | 221 | 15 | 14.73 | 0.24 |
| l13 | Long | Running in cities | 9 | 43 | 238 | 14 | 17.00 | 0.21 |
| l14 | Long | Lost keys | 10 | 33 | 211 | 14 | 15.07 | 0.30 |
| l15 | Long | Farmers market | 11 | 39 | 234 | 15 | 15.60 | 0.28 |
| l16 | Long | The famous journalist | 9 | 41 | 261 | 15 | 17.400 | 0.220 |
| l17 | Long | Too much drama | 7 | 39 | 237 | 14 | 16.929 | 0.179 |
| l18 | Long | People washing | 7 | 38 | 244 | 14 | 17.429 | 0.184 |
| s1 | Short | Fossil under construction | 5 | 20 | 119 | 7 | 17.00 | 0.25 |
| s2 | Short | The pottery shop | 5 | 20 | 116 | 7 | 16.57 | 0.25 |
| S3 | Short | A well architected rescue | 4 | 16 | 112 | 7 | 16.00 | 0.25 |
| s4 | Short | Sleeping aid | 5 | 18 | 117 | 7 | 16.71 | 0.28 |
| s5 | Short | The shy dancer | 4 | 18 | 112 | 7 | 16.00 | 0.22 |
| s5 | Short | The shoeshiner | 5 | 21 | 120 | 7 | 17.14 | 0.24 |
| s7 | Short | Boat life | 4 | 23 | 121 | 7 | 17.29 | 0.17 |
| s8 | Short | The book doctor | 4 | 18 | 117 | 7 | 16.71 | 0.22 |
| s9 | Short | Werewolves | 5 | 20 | 114 | 7 | 16.29 | 0.25 |
| s10 | Short | Shopping lists | 6 | 20 | 112 | 7 | 16.00 | 0.30 |
| s11 | Short | Family tree | 5 | 20 | 126 | 7 | 18.00 | 0.25 |
| s12 | Short | The shepherd | 4 | 20 | 120 | 6 | 20.00 | 0.20 |
| s13 | Short | Submarine | 6 | 22 | 123 | 7 | 17.571 | 0.273 |
| s14 | Short | Sandcastles | 5 | 20 | 126 | 7 | 18.000 | 0.250 |
| s15 | Short | The subway doctor | 4 | 17 | 122 | 7 | 17.429 | 0.235 |
| s16 | Short | Heart of stone | 4 | 22 | 123 | 7 | 17.571 | 0.182 |
| s17 | Short | The law on apples | 4 | 20 | 123 | 7 | 17.571 | 0.200 |
| s18 | Short | Overnight train | 6 | 18 | 126 | 7 | 18.000 | 0.333 |

### Supplementary Table S2: Schedule of ASRT stories administered during remote assessments.

*Tasks with delayed recall after distraction task (category or verbal fluency).

| Testing schedule | Remote Assessment Day | | | | | | | |
| --- | --- | --- | --- | --- | --- | --- | --- | --- |
|  | 1 | 2 | 3 | 4 | 5 | 6 | 7 | 8 |
| Schedule 1: ASRT stories administered (prior to February 27th) | l1  l2  l3 | l4  l5  l6 | l7  l8  l9 | l10*  l11*  l12* | l13*  l14*  l15* | s1  s2  s3 | s4  s5  s6 | - |
| Schedule 2: ASRTs administered  (after February 27th). | l1  l2  l3 | s1  s2  s3 | s4  s5  s6 | s7*  s8*  s9* | s10*  s11*  s12* | l10*  l11*  l12* | l13*  l14*  l15* | l4  l5  l6 |

### Supplementary Table S3: Daily State questionnaires administered

We’ll now ask you some questions on how you’ve been feeling today while completing the assessments, compared to how you’re usually feeling. Are you ready?

- I’m ready.

[Mood]

How has your mood been today, compared to how you usually feel?

- Much worse than usual.
- Worse than usual.
- Slightly worse than usual.
- The same as usual.
- Slightly better than usual.
- Better than usual.
- Much better than usual.

[Sleep]

How much did you sleep last night, compared to how much you usually sleep?

- Much less than usual.
- Less than usual.
- Somewhat less than usual.
- The same as usual.
- Somewhat more than usual.
- More than usual.
- Much more than usual.

[Attention]

How much did your mind wander while completing the assessments today, compared to how much it usually wanders?

- Much less than usual.
- Less than usual.
- Somewhat less than usual.
- The same as usual.
- Somewhat more than usual.
- More than usual.
- Much more than usual.

[Effort]

How hard did you try to do your best today while completing the assessments, compared to how much effort you usually put in?

- Much less than usual.
- Less than usual.
- Somewhat less than usual.
- The same as usual.
- Somewhat more than usual.
- More than usual.
- Much more than usual.

### Supplementary Table S4: Usability questionnaire administered on day 2 of old schedule and day 5 of revised schedule

Did you encounter technical problems while using this application to complete the challenges?

- Yes
- No

Did these technical problems prevent you from completing any of the challenges?

- Yes
- No

How easy did you find using this application on a scale of 1 to 5?

- 1 - Very hard to use
- 2
- 3
- 4
- 5 - Very easy to use

How interesting did you find completing this challenges on a scale of 1 to 5?

- 1 - Absolutely not interesting
- 2
- 3
- 4
- 5 - Very interesting

### Supplementary Figure SF 1: histograms of individual long stories (l1 to l3)

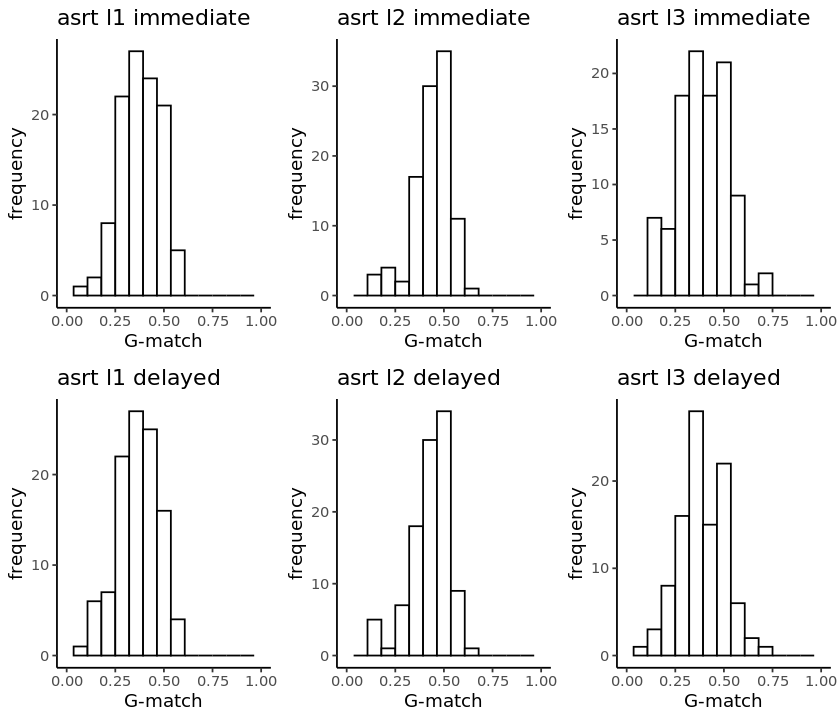

### Supplementary Figure SF 2: histograms of mean performance over long triplet (l1 to l3)

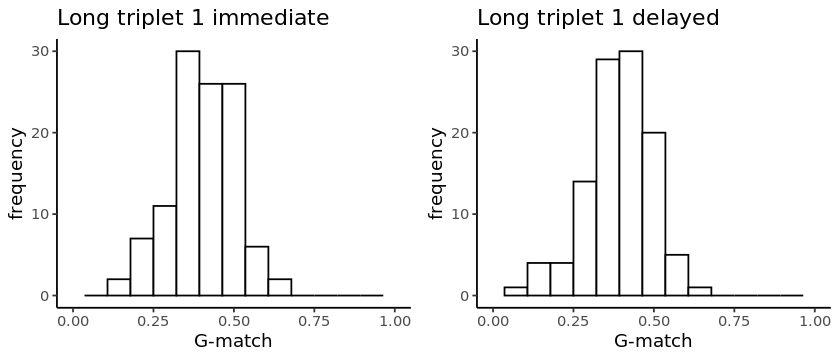

### Supplementary Figure SF 3: histograms of individual short stories (s1 to s3)

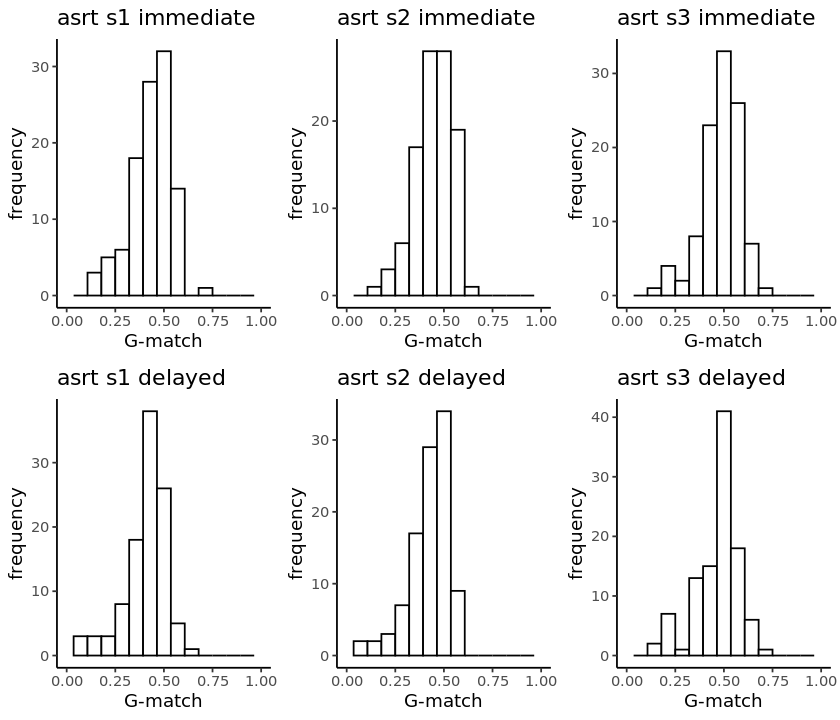

### Supplementary Figure SF 4: histograms of mean performance over short triplet (s1 to s3)

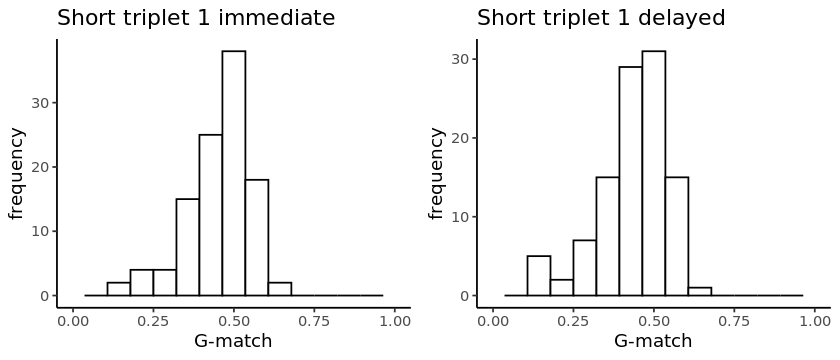

### Supplementary Table S5: task performance characteristics for long stories

Story recall metrics, means and standard deviations (sd) of number of words and recall accuracy as measured by G-match on immediate presentation and after a delay in MCI/Mild AD and Cognitively unimpaired group. The number of participants contributing to each mean is also shown (N).

| **Story ID** | **Story name** | **MCI/Mild AD** | | | | | | | | | | **Cognitively Unimpaired** | | | | | | | | | |
| --- | --- | --- | --- | --- | --- | --- | --- | --- | --- | --- | --- | --- | --- | --- | --- | --- | --- | --- | --- | --- | --- |
|  |  | **Immediate** | | | | | **Delayed** | | | | | **Immediate** | | | | | **Delayed** | | | | |
|  |  |  | **Number of words** | | **G-match** | |  | **Number of words** | | **G-match** | |  | **Number of words** | | **G-match** | |  | **Number of words** | | **G-match** | |
|  |  | **N** | mean | **sd** | mean | **sd** | **N** | mean | **sd** | **mean** | **sd** | N | mean | **sd** | **mean** | **sd** | N | mean | **sd** | **mean** | **sd** |
| l1 | No ordinary thief | 49 | 109.37 | 52.90 | 0.31 | 0.08 | 48 | 109.00 | 59.31 | 0.30 | 0.09 | 61 | 168.38 | 47.67 | 0.43 | 0.09 | 60 | 167.68 | 50.71 | 0.42 | 0.08 |
| l2 | The music lover | 45 | 157.51 | 67.72 | 0.38 | 0.10 | 48 | 152.10 | 80.95 | 0.36 | 0.11 | 58 | 210.03 | 49.77 | 0.48 | 0.07 | 58 | 215.40 | 53.69 | 0.47 | 0.06 |
| l3 | The walled city | 48 | 95.44 | 57.96 | 0.31 | 0.10 | 46 | 102.98 | 63.23 | 0.31 | 0.10 | 56 | 165.43 | 54.86 | 0.46 | 0.10 | 56 | 177.20 | 65.83 | 0.45 | 0.10 |
| l4 | The art competition | 43 | 102.81 | 74.14 | 0.26 | 0.10 | 38 | 103.68 | 79.01 | 0.26 | 0.10 | 58 | 170.88 | 77.56 | 0.34 | 0.08 | 58 | 170.93 | 77.20 | 0.34 | 0.07 |
| l5 | The Big Beetle | 41 | 95.00 | 59.50 | 0.28 | 0.10 | 37 | 94.81 | 61.66 | 0.27 | 0.11 | 58 | 155.48 | 52.54 | 0.38 | 0.08 | 55 | 161.89 | 59.80 | 0.37 | 0.07 |
| l6 | A soldier returns | 39 | 126.41 | 66.24 | 0.34 | 0.10 | 36 | 139.03 | 74.18 | 0.35 | 0.09 | 57 | 203.07 | 61.15 | 0.43 | 0.06 | 55 | 207.20 | 82.38 | 0.41 | 0.08 |
| l7 | The voice of God | 14 | 112.43 | 76.86 | 0.30 | 0.09 | 13 | 108.38 | 89.77 | 0.26 | 0.12 | 32 | 196.88 | 67.76 | 0.41 | 0.06 | 31 | 184.03 | 84.53 | 0.37 | 0.10 |
| l8 | The silent nun | 14 | 127.14 | 65.96 | 0.32 | 0.10 | 13 | 109.54 | 58.38 | 0.30 | 0.12 | 32 | 203.56 | 65.35 | 0.42 | 0.09 | 32 | 195.88 | 77.42 | 0.40 | 0.08 |
| l9 | The king and the doctor | 14 | 140.07 | 71.73 | 0.34 | 0.10 | 13 | 128.38 | 82.68 | 0.31 | 0.15 | 31 | 212.32 | 50.93 | 0.44 | 0.07 | 32 | 199.59 | 53.85 | 0.42 | 0.06 |
| l10 | Lost on an Island | 54 | 140.76 | 75.58 | 0.33 | 0.09 | 50 | 141.44 | 86.27 | 0.32 | 0.10 | 63 | 221.22 | 67.99 | 0.43 | 0.06 | 60 | 219.92 | 81.13 | 0.41 | 0.06 |
| l11 | Treasure of the Pyramids | 48 | 123.17 | 61.68 | 0.35 | 0.10 | 50 | 115.12 | 68.80 | 0.33 | 0.11 | 63 | 162.68 | 45.31 | 0.45 | 0.07 | 59 | 166.66 | 54.58 | 0.43 | 0.06 |
| l12 | The dry swimmer | 53 | 130.40 | 59.86 | 0.34 | 0.11 | 50 | 129.68 | 71.48 | 0.32 | 0.12 | 63 | 191.08 | 50.97 | 0.44 | 0.06 | 59 | 199.98 | 53.46 | 0.43 | 0.06 |
| l13 | Running in cities | 43 | 157.53 | 82.78 | 0.30 | 0.08 | 40 | 158.95 | 83.30 | 0.30 | 0.09 | 57 | 223.11 | 62.82 | 0.39 | 0.05 | 56 | 225.96 | 72.41 | 0.38 | 0.05 |
| l14 | Lost keys | 42 | 131.31 | 68.64 | 0.33 | 0.11 | 39 | 133.79 | 65.78 | 0.33 | 0.10 | 56 | 191.88 | 62.92 | 0.43 | 0.07 | 56 | 196.16 | 70.44 | 0.42 | 0.08 |
| l15 | Farmers market | 43 | 143.67 | 69.43 | 0.38 | 0.12 | 40 | 152.68 | 78.04 | 0.37 | 0.13 | 57 | 204.61 | 47.57 | 0.49 | 0.06 | 55 | 211.22 | 59.04 | 0.48 | 0.06 |

### Supplementary Table S6: task performance characteristics for short stories

Story recall metrics, means and standard deviations (sd) of number of words and recall accuracy as measured by G-match on immediate presentation and after a delay in MCI/Mild AD and Cognitively unimpaired group. The number of participants contributing to each mean is also shown (N).

| **Story ID** | **Story name** | **MCI/Mild AD** | | | | | | | | | | **Cognitively Unimpaired** | | | | | | | | | |
| --- | --- | --- | --- | --- | --- | --- | --- | --- | --- | --- | --- | --- | --- | --- | --- | --- | --- | --- | --- | --- | --- |
|  |  | **Immediate** | | | | | **Delayed** | | | | | **Immediate** | | | | | **Delayed** | | | | |
|  |  |  | **Number of words** | | **G-match** | |  | **Number of words** | | **G-match** | |  | **Number of words** | | **G-match** | |  | **Number of words** | | **G-match** | |
|  |  | **N** | mean | **sd** | mean | **sd** | **N** | mean | **sd** | mean | **sd** | N | mean | **sd** | **mean** | **sd** | **N** | **mean** | **sd** | **mean** | **sd** |
| s1 | Fossil under construction | 49 | 70.41 | 37.08 | 0.37 | 0.11 | 48 | 69.44 | 42.81 | 0.34 | 0.11 | 58 | 102.79 | 27.30 | 0.49 | 0.07 | 57 | 104.35 | 26.57 | 0.46 | 0.06 |
| s2 | The pottery shop | 47 | 75.13 | 37.30 | 0.40 | 0.11 | 46 | 75.02 | 44.96 | 0.37 | 0.11 | 56 | 101.48 | 25.92 | 0.49 | 0.07 | 57 | 108.58 | 32.84 | 0.47 | 0.06 |
| s3 | A well architected rescue | 48 | 81.08 | 37.30 | 0.43 | 0.11 | 47 | 73.49 | 48.22 | 0.41 | 0.13 | 57 | 100.33 | 26.20 | 0.53 | 0.07 | 57 | 103.44 | 24.42 | 0.51 | 0.07 |
| s4 | Sleeping aid | 42 | 83.05 | 37.96 | 0.42 | 0.09 | 40 | 80.05 | 52.19 | 0.36 | 0.12 | 61 | 109.98 | 37.11 | 0.50 | 0.08 | 59 | 119.46 | 43.80 | 0.50 | 0.08 |
| s5 | The shy dancer | 42 | 91.19 | 42.04 | 0.45 | 0.09 | 41 | 93.54 | 44.84 | 0.44 | 0.10 | 61 | 121.25 | 31.32 | 0.54 | 0.07 | 59 | 129.63 | 39.30 | 0.53 | 0.07 |
| s6 | The shoeshiner | 41 | 101.20 | 41.17 | 0.41 | 0.10 | 40 | 114.45 | 54.39 | 0.42 | 0.09 | 59 | 130.12 | 36.84 | 0.49 | 0.08 | 59 | 141.75 | 39.90 | 0.50 | 0.06 |
| s7 | Boat life | 31 | 68.39 | 41.80 | 0.35 | 0.09 | 31 | 64.77 | 42.22 | 0.32 | 0.11 | 32 | 108.22 | 42.22 | 0.45 | 0.08 | 31 | 113.35 | 38.76 | 0.44 | 0.07 |
| s8 | The book doctor | 32 | 79.13 | 37.06 | 0.36 | 0.08 | 30 | 81.37 | 36.24 | 0.36 | 0.08 | 31 | 114.52 | 38.86 | 0.45 | 0.07 | 31 | 123.16 | 41.76 | 0.43 | 0.06 |
| s9 | Werewolves | 31 | 72.35 | 25.74 | 0.41 | 0.11 | 29 | 72.69 | 35.06 | 0.40 | 0.10 | 32 | 103.56 | 29.83 | 0.52 | 0.08 | 31 | 110.77 | 38.82 | 0.50 | 0.07 |
| s10 | Shopping lists | 33 | 71.33 | 29.48 | 0.33 | 0.09 | 32 | 74.22 | 41.53 | 0.32 | 0.10 | 28 | 112.86 | 34.64 | 0.45 | 0.06 | 24 | 122.75 | 42.88 | 0.43 | 0.07 |
| s11 | Family tree | 33 | 78.64 | 39.68 | 0.38 | 0.13 | 32 | 88.38 | 50.57 | 0.38 | 0.12 | 26 | 137.08 | 33.56 | 0.53 | 0.09 | 22 | 159.73 | 52.95 | 0.51 | 0.08 |
| s12 | The shepherd | 32 | 77.81 | 34.67 | 0.37 | 0.08 | 32 | 87.28 | 39.33 | 0.38 | 0.08 | 25 | 129.40 | 41.75 | 0.47 | 0.05 | 22 | 145.09 | 53.75 | 0.44 | 0.04 |

### Supplementary Table S7: task performance characteristics for long and short triplets

Story recall metrics, means and standard deviations (sd) of number of words and recall accuracy as measured by G-match on immediate presentation and after a delay in MCI/Mild AD and Cognitively unimpaired group. The number of participants contributing to each mean is also shown (N).

| **Triplet ID** | **ASRTs included** | **Story length** | **MCI/Mild AD** | | | | | | | | | | **Cognitively Unimpaired** | | | | | | | | | |
| --- | --- | --- | --- | --- | --- | --- | --- | --- | --- | --- | --- | --- | --- | --- | --- | --- | --- | --- | --- | --- | --- | --- |
|  |  |  | **Immediate** | | | | | **Delayed** | | | | | **Immediate** | | | | | **Delayed** | | | | |
|  |  |  |  | **Number of words** | | **G-match** | |  | **Number of words** | | **G-match** | |  | **Number of words** | | **G-match** | |  | **Number of words** | | **G-match** | |
|  |  |  | **N** | mean | **sd** | mean | **sd** | **N** | mean | **sd** | **mean** | **sd** | N | mean | **sd** | **mean** | **sd** | N | mean | **sd** | **mean** | **sd** |
| Triplet L1 | l1, l2, l3 | Long | 49 | 119.75 | 53.27 | 0.33 | 0.09 | 48 | 121.48 | 62.48 | 0.32 | 0.09 | 61 | 180.70 | 46.53 | 0.46 | 0.08 | 60 | 186.48 | 51.62 | 0.45 | 0.07 |
| Triplet L2 | l4, l5, l6 | Long | 43 | 104.34 | 63.40 | 0.29 | 0.10 | 39 | 109.39 | 70.26 | 0.29 | 0.10 | 58 | 175.68 | 60.04 | 0.38 | 0.07 | 58 | 179.83 | 65.72 | 0.37 | 0.07 |
| Triplet L3 | l7, l8, l9 | Long | 14 | 126.55 | 66.85 | 0.32 | 0.09 | 14 | 120.64 | 70.43 | 0.29 | 0.11 | 32 | 203.16 | 56.97 | 0.42 | 0.06 | 32 | 192.61 | 66.55 | 0.40 | 0.07 |
| Triplet L4 | l10, l11, l12 | Long | 54 | 130.97 | 62.22 | 0.34 | 0.10 | 51 | 127.33 | 68.89 | 0.32 | 0.10 | 63 | 191.66 | 50.85 | 0.44 | 0.06 | 60 | 195.55 | 57.36 | 0.42 | 0.06 |
| Triplet L5 | l13, l14, l15 | Long | 43 | 145.21 | 67.76 | 0.34 | 0.10 | 40 | 147.40 | 71.79 | 0.33 | 0.10 | 57 | 206.63 | 52.25 | 0.44 | 0.06 | 57 | 211.47 | 61.18 | 0.42 | 0.06 |
| Triplet S1 | s1, s2, s3 | Short | 50 | 73.77 | 35.81 | 0.39 | 0.10 | 48 | 71.90 | 41.73 | 0.37 | 0.11 | 58 | 101.79 | 22.13 | 0.50 | 0.06 | 57 | 105.46 | 24.23 | 0.48 | 0.06 |
| Triplet S2 | s4, s5, s6 | Short | 42 | 91.58 | 35.18 | 0.43 | 0.08 | 41 | 94.96 | 46.25 | 0.41 | 0.09 | 63 | 119.79 | 31.24 | 0.51 | 0.06 | 59 | 130.28 | 37.08 | 0.51 | 0.06 |
| Triplet S3 | s7, s8, s9 | Short | 32 | 73.01 | 30.02 | 0.37 | 0.08 | 31 | 71.34 | 34.65 | 0.35 | 0.09 | 32 | 108.68 | 32.38 | 0.47 | 0.07 | 31 | 115.76 | 35.89 | 0.45 | 0.06 |
| Triplet S4 | s10, s11, s12 | Short | 34 | 74.80 | 31.81 | 0.36 | 0.10 | 32 | 83.29 | 41.18 | 0.36 | 0.09 | 28 | 125.52 | 31.06 | 0.48 | 0.06 | 24 | 139.83 | 45.38 | 0.46 | 0.06 |

### Supplementary Figure SF 5: Heatmap of ASRT parallel forms correlations for delayed recall and convergent validity with logical memory delayed recall, CDR-SB and PACC5 in the full sample

(Pairwise correlations coefficients for ASRTs reflect parallel task performance metrics for between n=72-108 participants, and with ASRTs and other other cognitive tests for n=89-108, depending on adherence patterns)

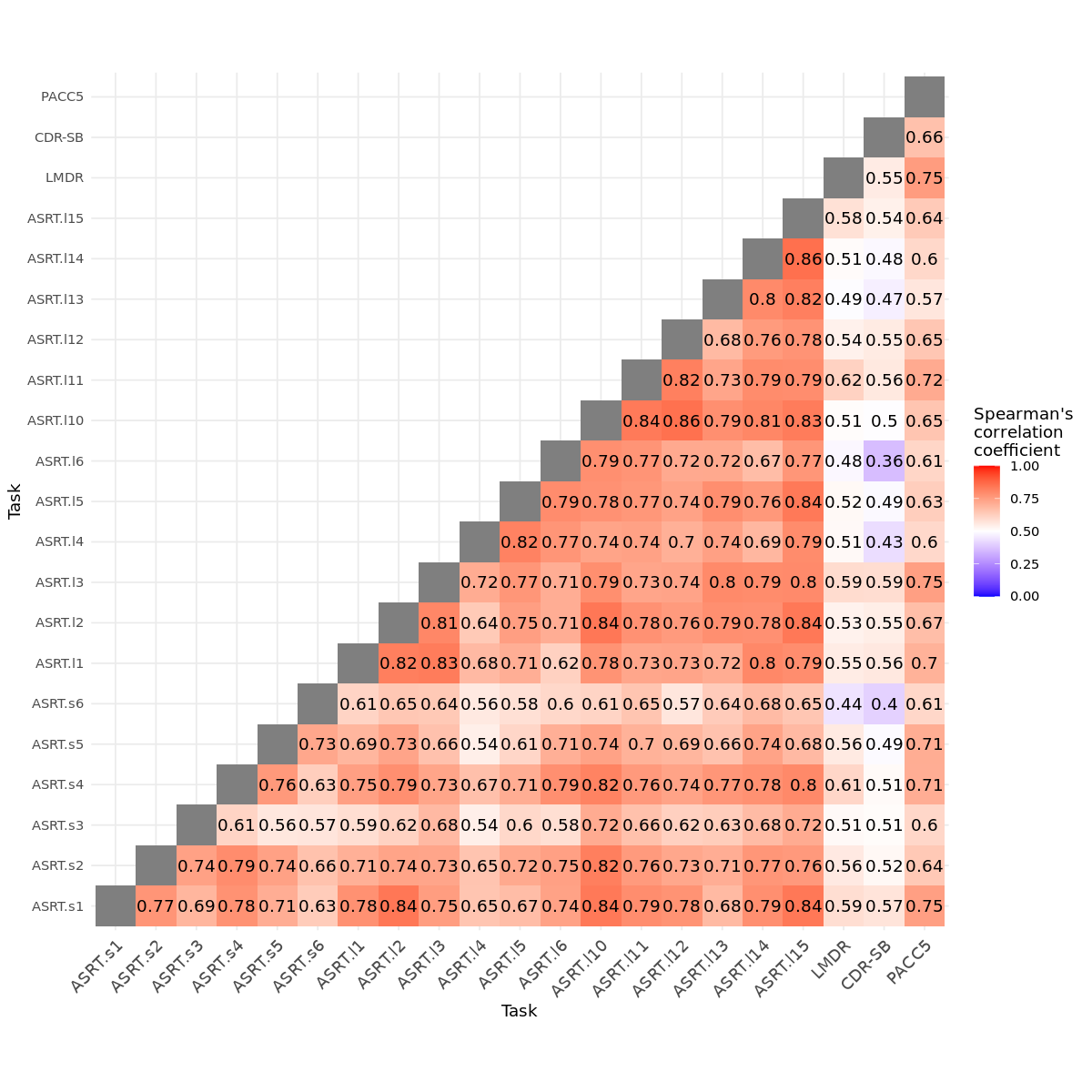

### Supplementary Figure SF 6: Heatmap of ASRT parallel forms correlations in MCI group only: immediate recall and convergent validity with logical memory immediate recall, CDR-SB and PACC5

(Pairwise correlations coefficients for ASRTs reflect parallel task performance metrics for between n=26-53 participants, and of ASRTs with other cognitive tests for n=35-53, depending on adherence patterns)

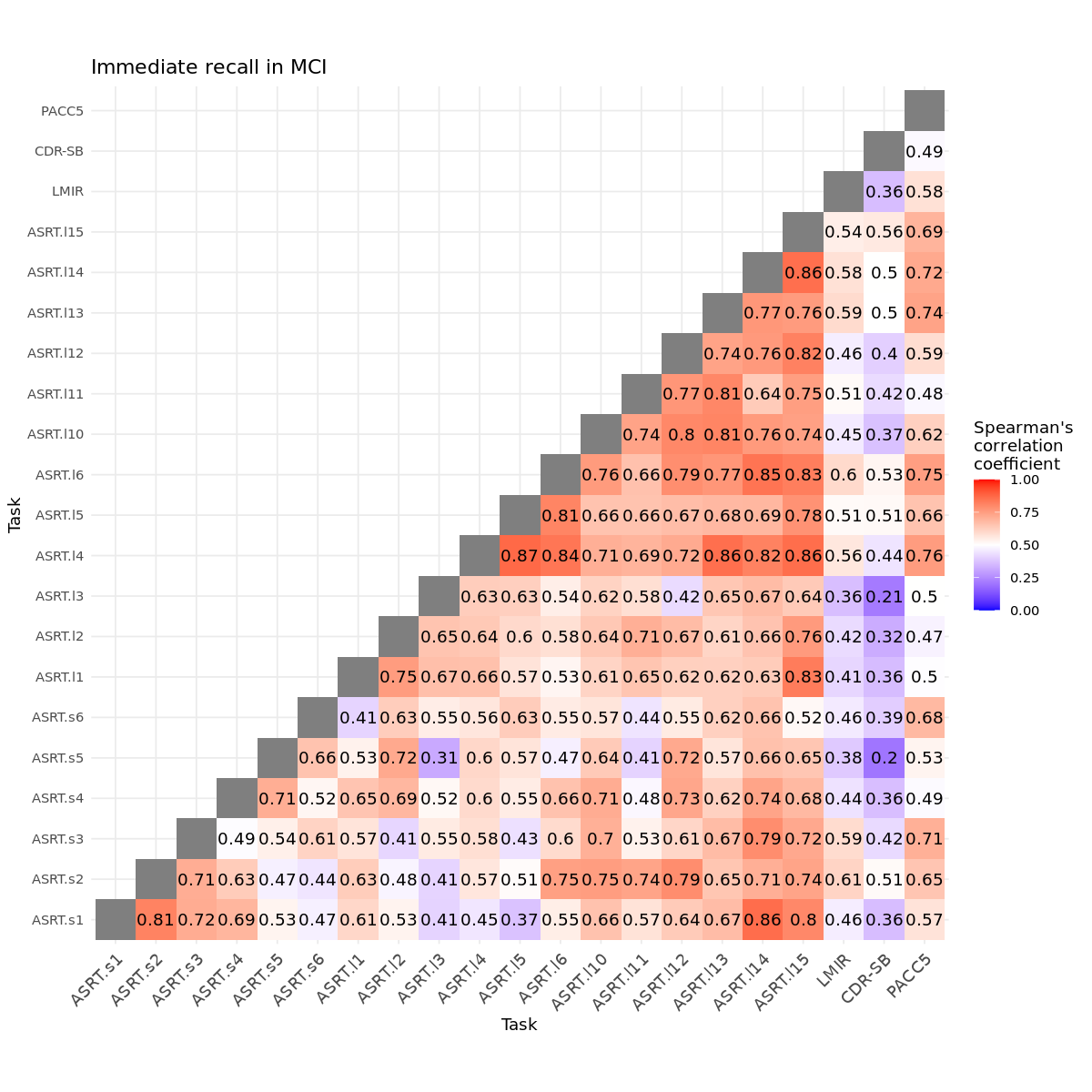

### Supplementary Figure SF 7: Heatmap of ASRT parallel forms correlations in MCI group only: delayed recall and convergent validity with logical memory immediate recall, CDR-SB and PACC5

(Pairwise correlations coefficients for ASRTs reflect parallel task performance metrics for between n=24-49 participants, and of ASRTs with other cognitive tests for n=34-49, depending on adherence patterns)

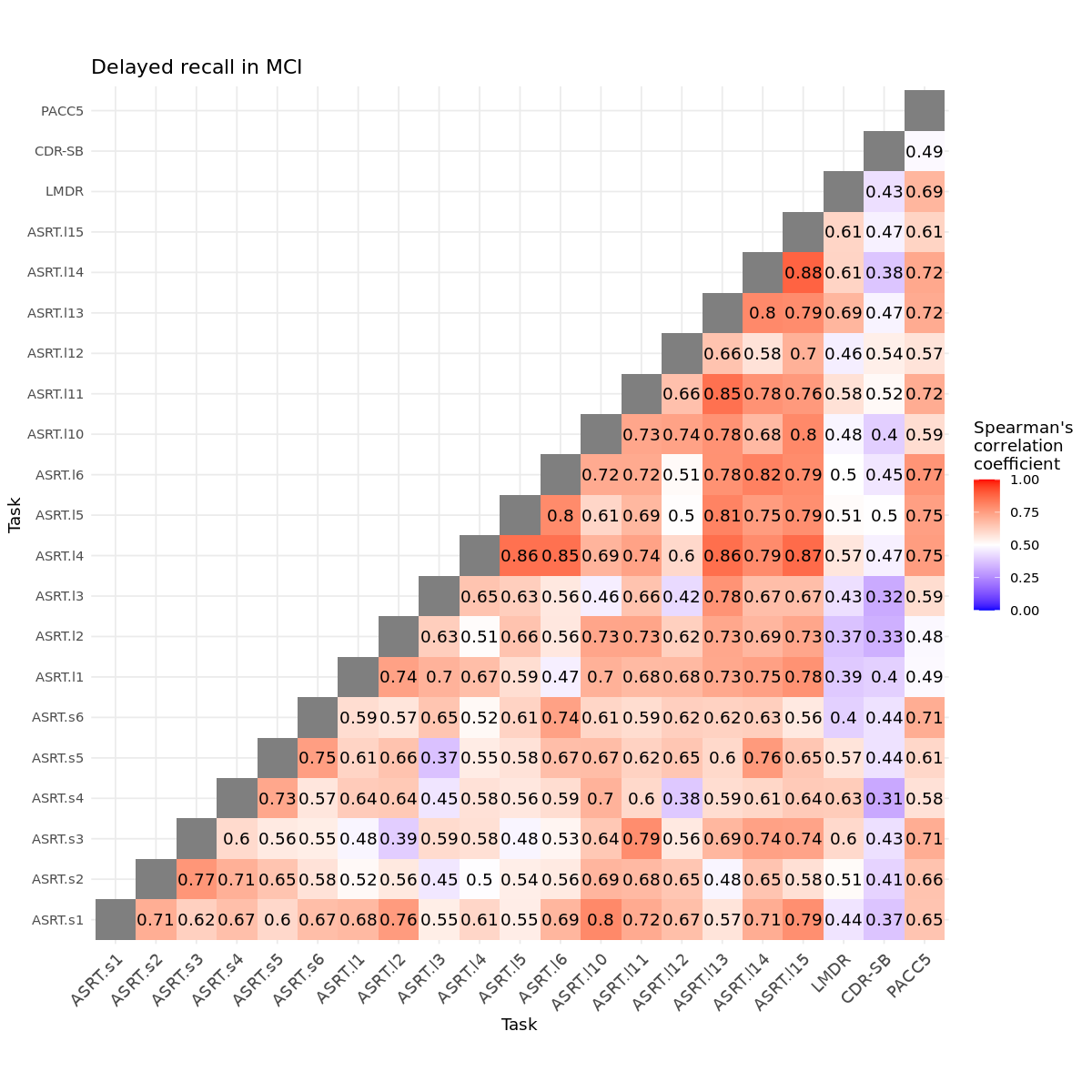

### Supplementary Figure SF 8: Heatmap of ASRT parallel forms correlations in CU group only: immediate recall and convergent validity with logical memory immediate recall, CDR-SB and PACC5

(Pairwise correlations coefficients for ASRTs reflect parallel task performance metrics for between n=46-63 participants, and of ASRTs with other cognitive tests for n=55-62, depending on adherence patterns)

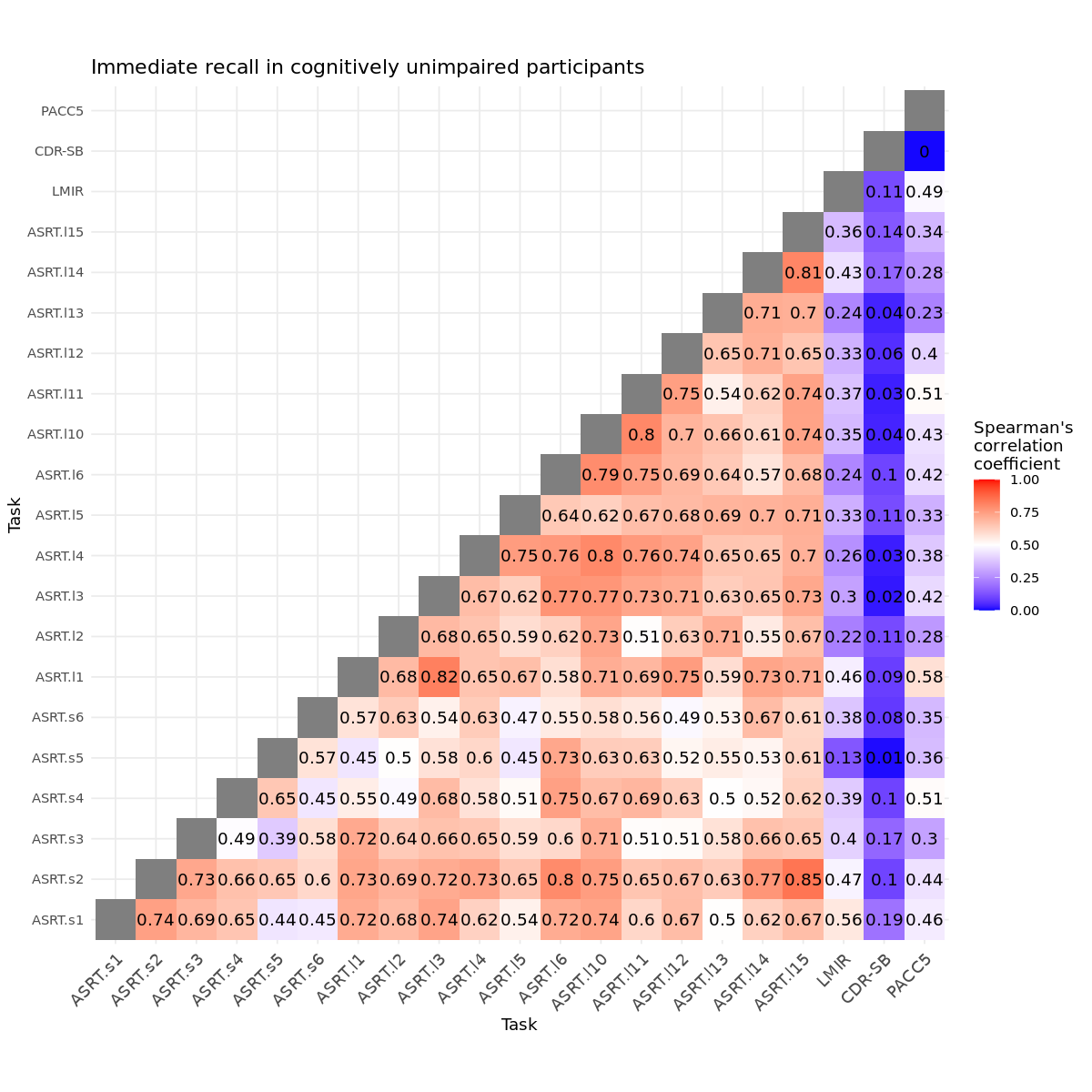

### Supplementary Figure SF 9: Heatmap of ASRT parallel forms correlations in CU group only: delayed recall and convergent validity with logical memory immediate recall, CDR-SB and PACC5

(Pairwise correlations coefficients for ASRTs reflect parallel task performance metrics for between n=46-59 participants, and of ASRTs with other cognitive tests for n=54-60, depending on adherence patterns)

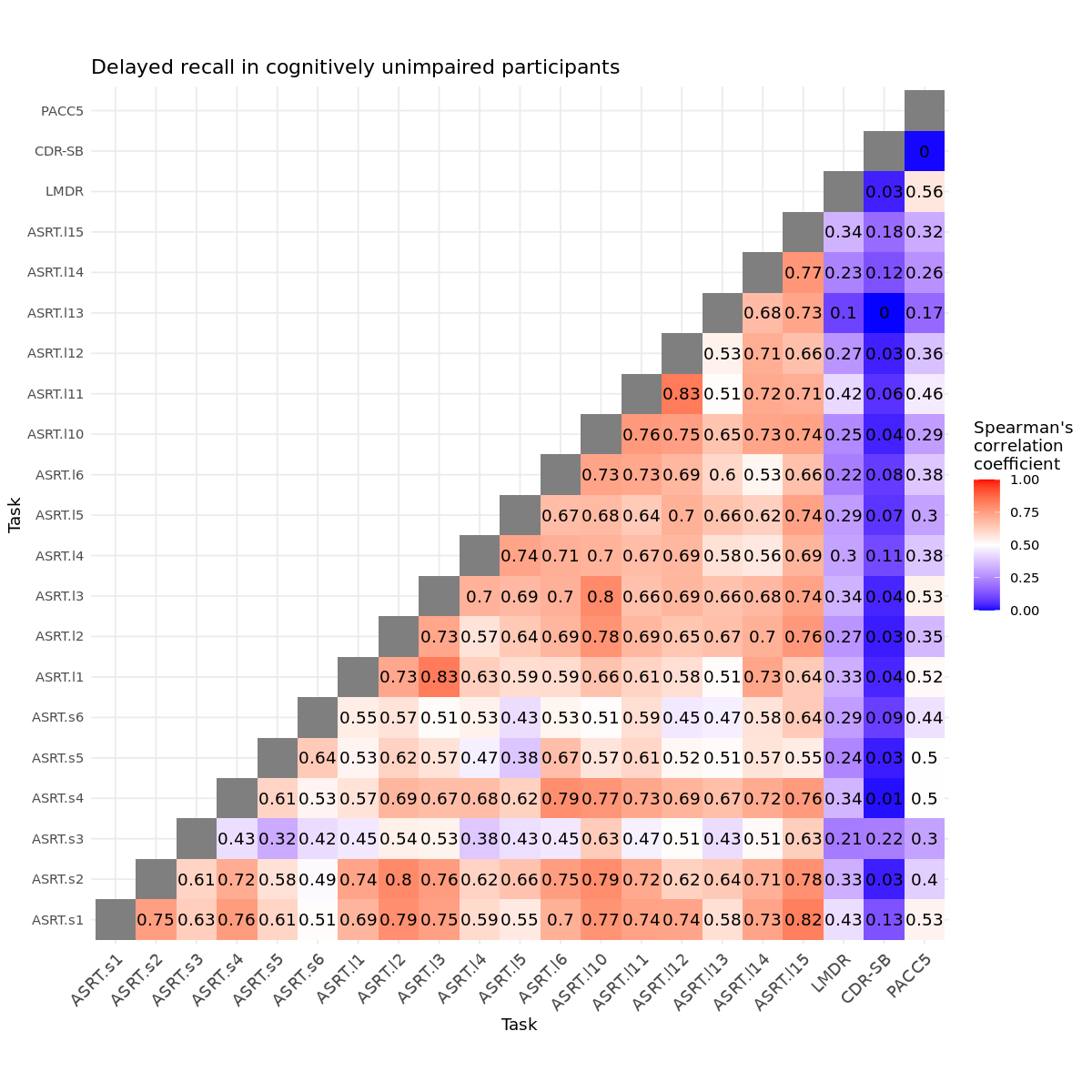

### Supplementary Figure SF 10: Heatmap of ASRT triplets in full sample:

A) immediate recall (pairwise comparisons for n=81-93 participants); B) delayed recall (pairwise comparisons for n=77-90 participants). Correlations coefficients for correlations with Logical memory immediate recall (LMIR) or delayed recall (LMDR), CDR-SB and PACC5 are also presented

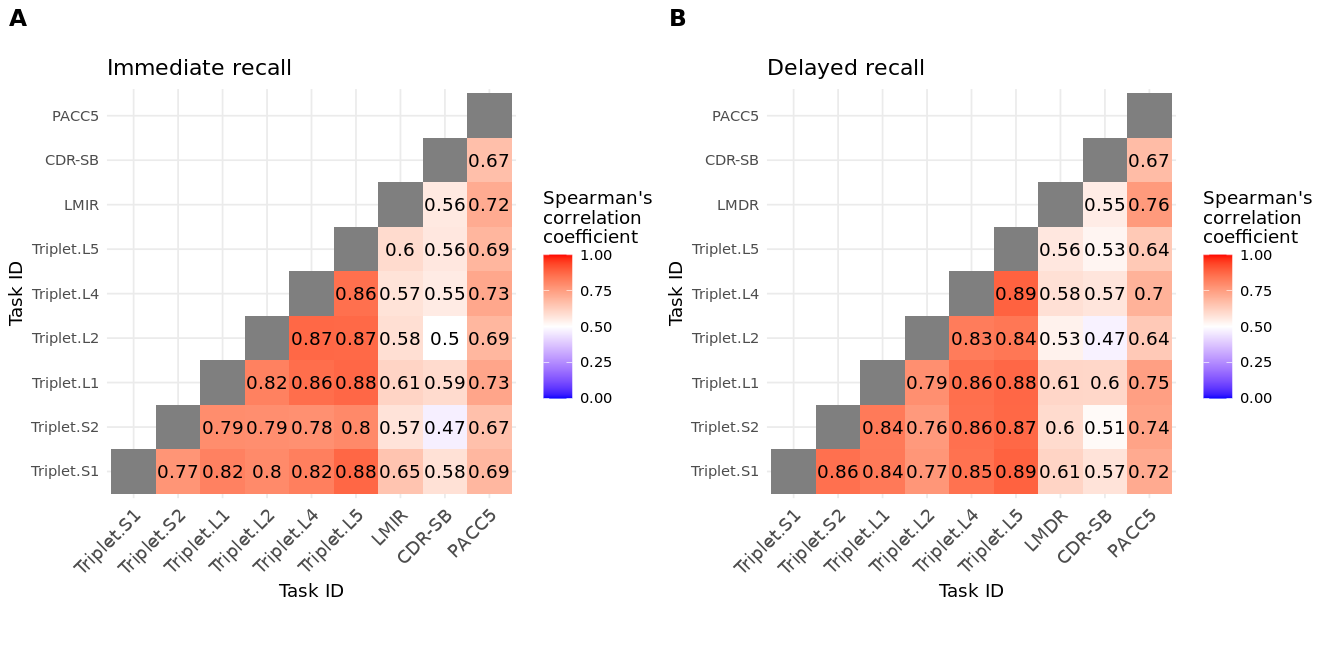

### Supplementary Figure SF 11: Heatmap of ASRT triplets in MCI subsample:

A) immediate recall (pairwise comparisons for n=27-38 participants); B) delayed recall (pairwise comparisons for n=26-35 participants). Correlations coefficients for correlations with Logical memory immediate recall (LMIR) or delayed recall (LMDR), CDR-SB and PACC5 are also presented

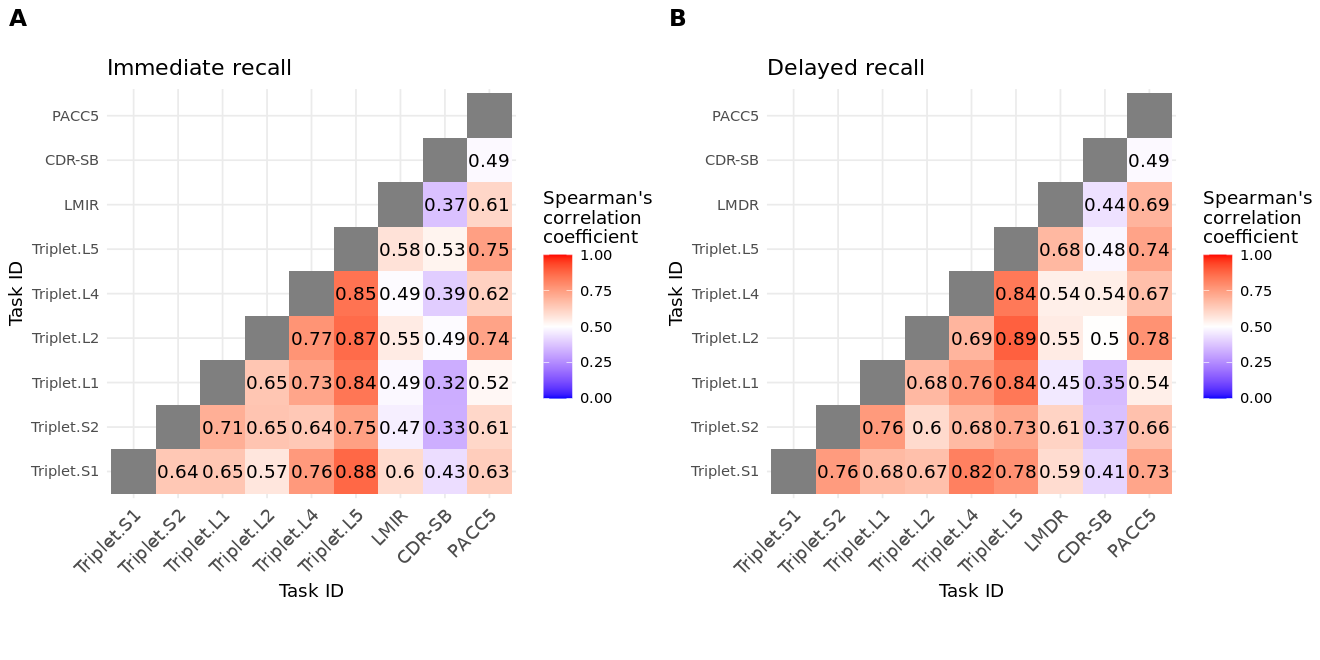

### Supplementary Figure SF 12: Heatmap of ASRT triplets in MCI subsample:

A) immediate recall (pairwise comparisons for n=49-55 participants); B) delayed recall (pairwise comparisons for n=48-53 participants). Correlations coefficients for correlations with Logical memory immediate recall (LMIR) or delayed recall (LMDR), CDR-SB and PACC5 are also presented.

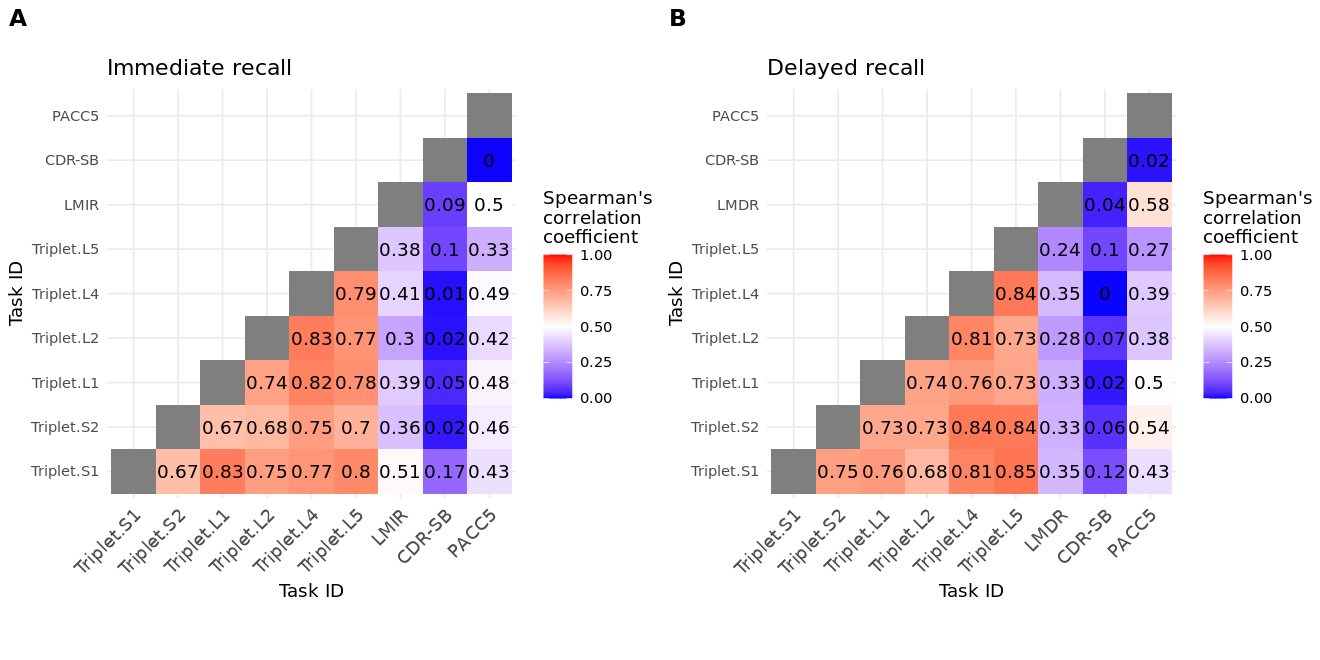
